# Clonal hematopoiesis and risk of chronic liver disease

**DOI:** 10.1101/2022.01.17.22269409

**Authors:** Waihay J. Wong, Connor Emdin, Alexander Bick, Seyedeh M. Zekavat, Abhishek Niroula, James Pirruccello, Laura Dichtel, Gabriel Griffin, Md Mesbah Uddin, Christopher J. Gibson, Veronica Kovalcik, Amy E. Lin, Marie E McConkey, Amelie Vromman, Rob S. Sellar, Peter G. Kim, Mridul Agrawal, Joshua Weinstock, Michelle T. Long, Bing Yu, Rajarshi Banerjee, Rowan C. Nicholls, Matt Kelly, Po-Ru Loh, Steve McCarroll, Eric Boerwinkle, Ramachandran Vasan, Siddhartha Jaiswal, Andrew Johnson, Raymond T. Chung, Kathleen Corey, Daniel Levy, Christie Ballantyne, NHLBI TOPMed Hematology Working Group, Benjamin L. Ebert, Pradeep Natarajan

## Abstract

Chronic liver disease is a major public health burden worldwide. Despite various liver injury mechanisms, progression of chronic liver disease follows a common pathway of liver inflammation, injury and fibrosis. We examined the association between clonal hematopoiesis of indeterminate potential (CHIP) and chronic liver disease in 58,358 individuals from four prospective cohorts with whole exome sequencing data (Framingham Heart Study, Atherosclerosis Risk in Communities Study, UK Biobank and Mass General Brigham Biobank). CHIP was associated with an increased risk of prevalent chronic liver disease (OR 2.70 CI 1.42, 5.16, p=0.002) and incident chronic liver disease (HR 2.01 CI 1.46, 2.79, p=0.001) from both alcoholic and nonalcoholic causes. Individuals with CHIP showed 75% greater odds of MRI detectable liver inflammation and fibrosis (5.9% versus 3.5%, p=0.007) compared to those without CHIP. To assess causality, Mendelian randomization analyses showed that genetic predisposition to CHIP was associated with a greater risk of chronic liver disease (OR 2.37 CI 1.57, 3.6, p<0.001). In a dietary model of nonalcoholic steatohepatitis (NASH), mice transplanted with *Tet2*-deficient hematopoietic cells demonstrated more severe liver inflammation and fibrosis. These effects were mediated via NLRP3 inflammasome and increased downstream inflammatory cytokine expression, including IL6. In summary, clonal hematopoiesis is associated with an elevated risk of liver inflammation and chronic liver disease progression via an aberrant inflammatory response.

## Introduction

Chronic liver disease affects up to 19% of Americans in an age-dependent fashion.(*1*) Chronic liver disease is characterized by an inflammatory and fibrotic response to an initial insult, most commonly steatosis from excess alcohol consumption, obesity, or viral hepatitis.(*2*) In all cases, the progression of chronic liver disease encompasses a spectrum of histopathologic changes from liver fat accumulation (steatosis) to liver inflammation and hepatocyte injury (steatohepatitis), fibrosis and cirrhosis. However, the factors that influence progression from steatosis to inflammation and fibrosis are poorly understood.

Liver inflammation and fibrosis are mediated in part by nonparenchymal cells of the liver, including sinusoidal endothelial cells, dendritic cells, lymphocytes, and macrophages. Resident liver macrophages (Kupffer cells) and bone marrow derived monocytes and macrophages have been implicated in responses to liver injury in both murine models and humans.(*3, 4*) In non-alcoholic fatty liver disease (NAFLD), macrophage recruitment is required for progression to non-alcoholic steatohepatitis (NASH), whereas inhibition of monocyte recruitment prevents disease progression in mouse models.(*5*–*7*) Furthermore, monocyte-derived inflammatory macrophages are enriched in liver samples from patients who progress from NASH to cirrhosis.(*5*)

Dysregulated inflammatory responses can occur in the setting of clonal hematopoiesis of indeterminate potential (CHIP), which is characterized by the expansion of hematopoietic cells bearing oncogenic somatic mutations most frequently in the genes *DNMT3A, TET2*, and *ASXL1*.(*8*) Whole exome sequence analysis of blood DNA has led to the recognition that CHIP is a common phenomenon with increasing prevalence in older age, present in greater than 10% of persons over the age of 70.(*9*–*11*) CHIP is associated with future risk of hematologic malignancy,(*9, 10*) all-cause mortality(*9, 10*), and atherosclerotic cardiovascular disease.(*12, 13*) Murine models have revealed the pro-inflammatory role of macrophages derived from mutant CHIP clones and their contributions to atherogenesis.(*12, 14, 15*) Given the pervasive nature of circulating immune cells, we hypothesized that CHIP could potentially influence the trajectory of steatohepatitis and cirrhosis through aberrant inflammation in the liver.

Here, we test the hypothesis that CHIP is a novel risk factor for chronic liver disease by (1) associating CHIP with independent risk of chronic liver disease in three distinct cohorts, (2) associating CHIP with subclinical advanced liver imaging biomarkers, (3) causal inference with human germline genetics, (4) causal inference in mouse models, and (5) mechanistic inference using mouse models and human germline genetics.

## Methods

### Study samples

We examined the association between CHIP and chronic liver disease using five datasets (**Supp. Figure 1**). For analysis of prevalent disease, we used data from Framingham Heart Study (FHS, n=4,114) and Atherosclerosis Risk in Communities study (ARIC, n=7,414, **Table 1**). For analysis of incident disease, we tested the association of CHIP and genotyped CHIP variants with incident chronic liver disease in distinct groups of individuals from the UK Biobank who underwent whole exome sequencing (n=201,409) or genotyping (n=239,316). Individuals in the UK Biobank who underwent WES were selected to enrich for participants for whom magnetic resonance imaging results and linked hospitalization and primary care records were available.(*16*) Individuals with leukemia or other hematologic malignancy at the time of DNA collection were excluded from analysis.

**Table 1.** Baseline characteristics of participants in samples analyzed. ARIC, Atherosclerosis Risk in Communities study; BMI, body-mass index; FHS, Framingham Heart Study; MGB, Mass General Brigham; SD, standard deviation; WES, whole exome sequencing.

| Characteristics | FHS, n=4,230 | ARIC, n=7,414 | UK Biobank WES, n=201,409 | UK Biobank Array, n=239,316 | MGB Biobank, n=1,482 |
| --- | --- | --- | --- | --- | --- |
| Age, mean years (SD) | 61.1 (15.7) | 57.4 (6.0) | 56.5 (8.1) | 57.0 (8.1) | 53 (12) |
| Women, n (%) | 2,292 (54%) | 4,358 (56%) | 110,192 (55%) | 127,600 (53%) | 871 (59%) |
| BMI, mean kg/m<sup>2</sup> (SD) | 27.8 (5.5) | 27.7 (5.4) | 27.4 (4.8) | 27.4 (4.8) | 871 (58%) |
| Current smoking, n (%) | 610 (15%) | 1,992 (26%) | 13,358 (7%) | 20,819 (9%) | NA |
| Alcohol intake, mean drinks per week (SD) | 6.8 (16.6) | 5.6 (8.8) | 7.9 (9.8) | 8.1 (10.3) | 4.4 (7.4) |
| History of diabetes mellitus, n (%) | 166 (4%) | 674 (9%) | 9,676 (5%) | 12,438 (5%) | 95 (7%) |
| CHIP prevalence, n (%) | 369 (9%) | 333 (4%) | 11,783 (5%) | 189 (0.1%) | 90 (6%) |
| CHIP<sub>≥10%</sub> prevalence, n (%) | 342 (8%) | 267 (4%) | 6,188 (3%) | 189 (0.1%) | 51 (4%) |

We defined chronic liver disease as the development of liver fibrosis or cirrhosis, combining ICD10 diagnostic codes that have been demonstrated to show high specificity for identifying patients with cirrhosis compared to physician review and which associate strongly with known genetic loci for cirrhosis (**Supp. Table 1**).(*17*) Alcohol-related liver disease was defined as chronic liver disease among individuals with excess alcohol intake (≥21 drinks per week for men or ≥14 drinks per week for women) and no history of hepatitis B or hepatitis C.(*18*) Non-alcoholic steatohepatitis (NASH) was defined as chronic liver disease among individuals consuming fewer than 21 drinks per week for men and fewer than 14 drinks per week for women and no history of hepatitis B or hepatitis C as outlined in the American Association for the Study of Liver Diseases guidelines.(*18*)

As the above analyses of chronic liver disease were based primarily on ICD10 codes, we also examined whether CHIP predisposes to biopsy-proven NASH (n=1,482). For this analysis, we analyzed 114 individuals with biopsy-proven NASH and 1368 age- and sex-matched control individuals in the Massachusetts General Brigham Biobank. CHIP was ascertained through whole exome sequencing of blood samples deposited at the time of enrollment.

### Whole-exome sequencing and CHIP ascertainment

We identified individuals with CHIP based on a prespecified list of variants in 74 genes that are recurrently mutated in myeloid cancers (**Supp. Table 2**). For prevalent liver disease analyses in FHS and ARIC, CHIP was ascertained using WES.(*19, 20*) For incident liver disease analysis in UK Biobank, CHIP was ascertained using whole exome sequencing in the subset of individuals for which exome sequencing data was available (n=201,409). For the UK Biobank subset in which genotyping array data was available (n=239,316), CHIP was ascertained using array-derived genotyped variants in *ASXL1, DNMT3A, JAK2*, and *TET2* (**Supp. Table 3**). We examined genotyping fidelity of each variant by manually examining imaging files with ScatterShot. Individuals included in WES and prevalence analysis were excluded from this analysis. In the primary analysis, we examined CHIP with a variant allele fraction ≥10%. We separately analyzed CHIP with a variant allele fraction below 10%.

### Mouse models

Lethally irradiated *Ldlr*^-/-^ and B6.SJL mice were transplanted with bone marrow cells from *vavCre*^+^*Tet2*^fl/-^ *vavCre*^+^*Tet2*^fl/*-*^*Nlrp3*^*-/-*^ and control *vavCre*^+^ mice. Transplanted mice were fed an atherogenic Western diet containing 0.2% cholesterol and 42 kcal % fat (TD.88137; Envigo) or a choline-deficient, L-amino acid defined, high-fat diet containing 60 kcal % fat and 0.1% methionine (CDAHFD; Research Diets). After a defined diet period, mice were sacrificed and perfused mouse livers were harvested and processed for histologic grading of steatohepatitis, quantification of fibrosis by image analysis, fluorescence-activated cell sorting of liver macrophages, RNA extraction and transcriptome analysis (see Extended Methods).

### Statistical analysis

We tested for the association of CHIP status with prevalent chronic liver disease using logistic regression, adjusting for age, sex, type 2 diabetes, and smoking. Additional analyses were performed after adjustment for alcohol consumption and body mass index. For incident analysis, we used Cox proportional hazards regression with adjustment for age, sex, type 2 diabetes, and smoking. Further adjustment for alcohol consumption and body mass index was performed as a sensitivity analysis. To aggregate the effect size of CHIP on liver disease across cohorts, inverse variance weighted fixed effects meta-analysis was performed. To test the association of CHIP status with liver imaging biomarkers, including liver fat (proton density fat fraction ≥ 5%) and liver inflammation and fibrosis (proton cT1 ≥ 795 ms), we used logistic regression with adjustment for age and sex. We used Mendelian randomization analysis with robust adjusted profile score (MR-RAPS) to examine the association of genetic predisposition to CHIP with cirrhosis risk, which provides for control of type 1 error rate when using sub-genome wide significant genetic variants.(*21*) Statistical analyses of mouse experiments are described in the Extended Methods. All statistical analyses were conducted using R version 3.5 or Graphpad Prism 8.3.1. Mendelian randomization analyses were conducted using the mr.raps R package.

## Results

### Association of CHIP with prevalent and incident chronic liver disease

We examined whether CHIP is associated with an elevated risk of chronic liver disease, ascertained using clinician interview and ICD codes (**Supp. Table 1**), in three cohorts. We tested the association of CHIP with prevalent chronic liver disease using data from the Framingham Heart Study (FHS, n=4,114) and Atherosclerosis Risk in Communities study (ARIC, n=7,414). We tested the association of CHIP with incident chronic liver disease using the subsample of the UK Biobank for which whole exome sequencing had been performed (n=201,409) as well as the subsample of the UK Biobank for which array genotyping was performed (n=239,316). In these cohorts, the mean (standard deviation) age range was 57-61 (6-16) years. The prevalence of CHIP, ascertained by exome sequencing, varied between 4% and 9% (**Table 1**). As in previous cohorts, *DNMT3A* and *TET2* were the most commonly mutated genes in CHIP (40%, **Supp. Figure 2**). The prevalence of CHIP increased across cohorts with increasing age (**Supp. Figure 3**). Known associations with CHIP, including age, sex, type 2 diabetes mellitus, smoking and self-reported ethnicity exhibited similar associations to CHIP across cohorts (**Supp. Table 3**). In the UK Biobank, CHIP variants ascertained through genotyping had high positive predictive value for exome sequencing-ascertained CHIP (90%), showed similar association with age to CHIP ascertained by exome sequencing (OR 1.04 per year, p<0.001, **Supp. Table 4**), and were strongly associated with incident myeloid hematologic malignancy (HR 106 CI 72, 158, p<0.001).

In FHS and ARIC, individuals with CHIP with a variant allele fraction (VAF) ≥ 10% (CHIP_≥10%_) were at increased odds of prevalent chronic liver disease after adjustment for age, sex, type 2 diabetes and smoking (OR 2.70 CI 1.42, 5.16, p=0.002 **Figure 1A**). In the UK Biobank, individuals with CHIP_>10%_ demonstrated increased risk for incident chronic liver disease (HR 1.82 CI 1.25, 2.65, p=0.001, respectively) (**Figure 1A**). Overall, CHIP_>10%_ was associated with a two-fold risk of prevalent or incident chronic liver disease (OR 2.01 CI 1.46, 2.79, p<0.001). No evidence of heterogeneity was observed between estimates among cohorts or estimates between prevalent and incident chronic liver disease. Individuals with CHIP continued to be at elevated risk of chronic liver disease when adjusted for baseline alcohol consumption, body mass index, alanine transaminase levels, aspartate transaminase levels and alkaline phosphatase levels (OR 2.11 CI 1.80, 2.47, p<0.001). When interaction terms between CHIP status, current smoking and alcohol consumption in weekly drinks were included in the model, there was no evidence of an interaction between current smoking (p interaction = 0.48) or alcohol consumption in weekly drinks (p interaction = 0.95).

**Figure 1.**
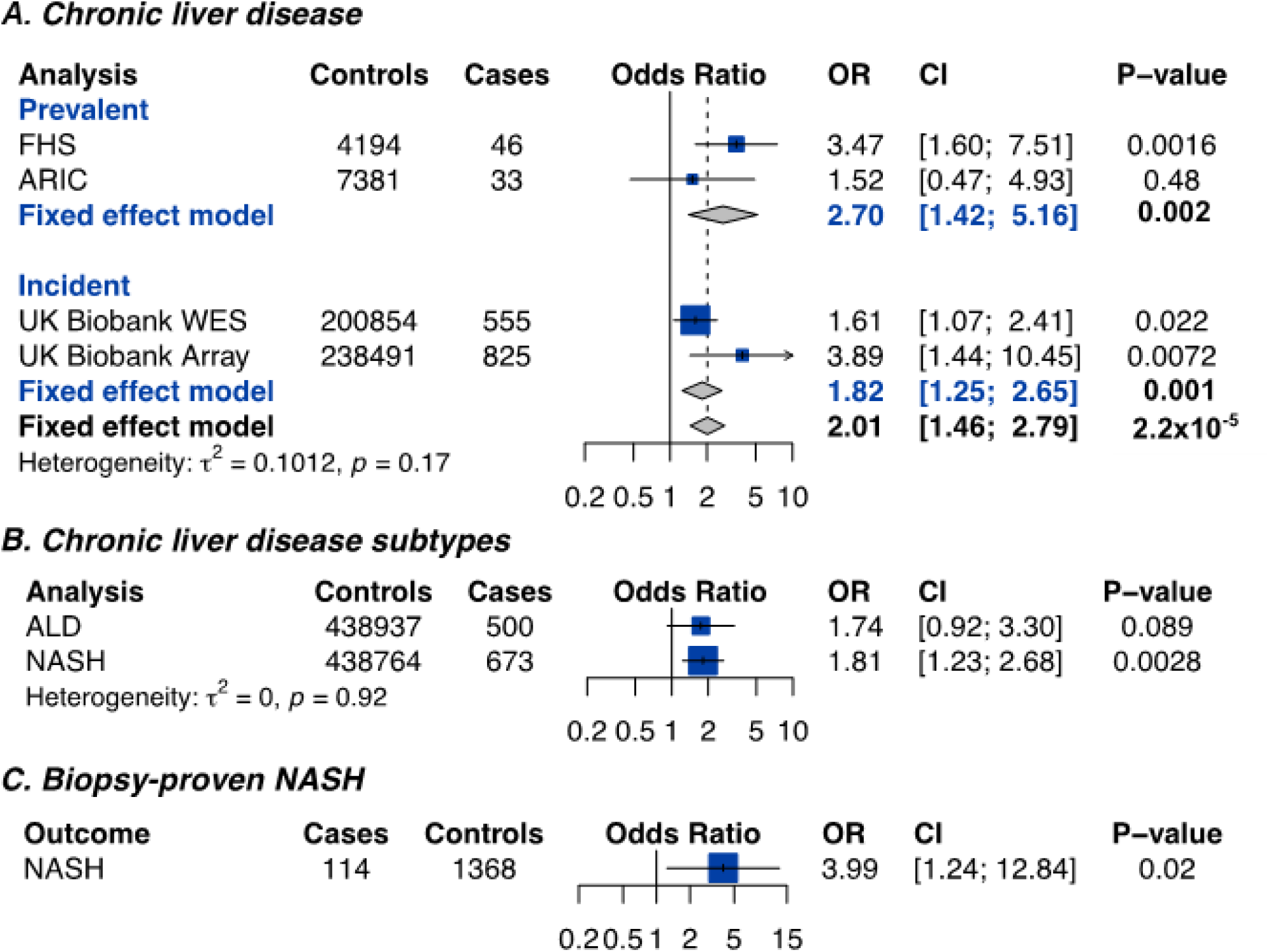
Clonal hematopoiesis of indeterminate potential is associated with chronic liver disease. **(A)** Association of clonal hematopoiesis of indeterminate potential with prevalent chronic liver disease. **(B)** Association of clonal hematopoiesis of indeterminate potential with incident chronic liver. **(C)** Association of clonal hematopoiesis with subtypes of incident chronic liver disease in the UK Biobank. Estimates in prevalent analyses were derived using logistic regression, with adjustment for age sex, type 2 diabetes and smoking. Estimates in incident analyses were derived using Cox proportional hazards regression, with adjustment for age, sex, type 2 diabetes, and smoking. NASH was defined as chronic liver disease among individuals with a body mass index of 30 kg/m^2^ or more who consumed 21 drinks or less per week for men and 14 drinks or less per week for women. Alcohol-related liver disease was defined as chronic liver disease among individuals who consumed 21 drinks or more for men or 14 drinks per week or more for women. ARIC, Atherosclerosis Risk in Communities study; CI, 95% confidence interval; FHS, Framingham Heart Study; OR, odds ratio; UKBB, UK Biobank.

CHIP with VAF <10% had a lower risk of chronic liver disease than CHIP with VAF ≥ 10% (OR 1.28 vs. 2.01, p_interaction_ = 0.15) (**Supp. Figure 4**). *JAK2*-mutant CHIP was associated with a highly elevated risk of chronic liver disease (OR 17.65 CI 4.32, 72.15, p<0.001), potentially due to established prothrombotic effects; nevertheless, non-*JAK2*-mutant CHIP was also associated with an elevated risk of chronic liver disease (OR 1.88 CI 1.09, 3.24, p=0.02) (**Supp. Figure 5**). *TET2*-mutant CHIP was independently associated with a highly elevated risk of chronic liver disease (OR 5.35 CI 2.22, 12.93, p<0.001). Autosomal chromosomal mosaicism of blood cells, a form of clonal hematopoiesis that is distinct from CHIP, was not associated with risk of chronic liver disease in the UK Biobank (OR 1.18 CI 0.97, 1.45, p=0.10).(*22*)

We next examined cumulative risk of chronic liver disease in the UK Biobank by array-derived CHIP status. Individuals without CHIP had a 1% cumulative incidence of chronic liver disease by age 80 years. In contrast, individuals with CHIP had a 6% cumulative incidence of liver disease (p<0.001, **Supp. Figure 6**). In comparison, individuals with morbid obesity (BMI > 35 kg/m^2^) without CHIP had a 2.5% cumulative incidence of liver disease.

When the subtypes of liver disease were examined, individuals with CHIP were at similarly elevated risk of both alcohol-related liver disease (ALD) and NASH (**Figure 1B**). Only seven individuals with virus-related chronic liver disease (chronic liver disease and a history of hepatitis C or hepatitis B) could be identified, preventing ascertainment of the association of CHIP with virus-related chronic liver disease. To further confirm the association of CHIP with NASH, we identified 114 individuals with biopsy-proven NASH and 1368 age- and sex-matched controls in the Mass General Brigham Biobank. Patients with biopsy-proven NASH were five times as likely to have CHIP as control subjects (OR 3.99 CI 1.24, 12.84, p=0.02, **Figure 1C)**.

### Association of CHIP with liver imaging and disease biomarkers

We studied whether CHIP may be associated with increased liver inflammation and fibrosis specifically using magnetic resonance imaging data from 8251 individuals in the UK Biobank. CHIP was associated with increased liver inflammation (proton cT1 ≥ 795 ms, OR 1.74 CI 1.16, 2.60, p= 0.007, **Figure 2A and 2B**). CHIP was not associated with hepatic steatosis (proton density fat fraction ≥ 5%, OR 0.98 CI 0.75, 1.28, p=0.89, **Figure 2A and 2B**). Using data from the UK Biobank, we also examined the relationship between CHIP and serum biomarkers in up to 393,128 individuals (**Supp. Figure 7**). CHIP was not associated with alanine transaminase levels or aspartate transaminase levels, but was associated with modest increases in alkaline phosphatase (0.015 U/L, p=0.004), total bilirubin (0.31 m/dl, p=0.0002) and C-reactive protein (0.19 mg/dl, p=0.03). CHIP was also associated with a modest elevation in platelet count (8.5 per uL, p<0.001) and leukocyte count (0.11 × 10^9^ cells/L, p=0.01). When *JAK2* CHIP was excluded, CHIP continued to be associated with elevations in alkaline phosphatase, total bilirubin and C-reactive protein, but was not associated with increased platelet count (**Supp. Figure 8**).

**Figure 2.**
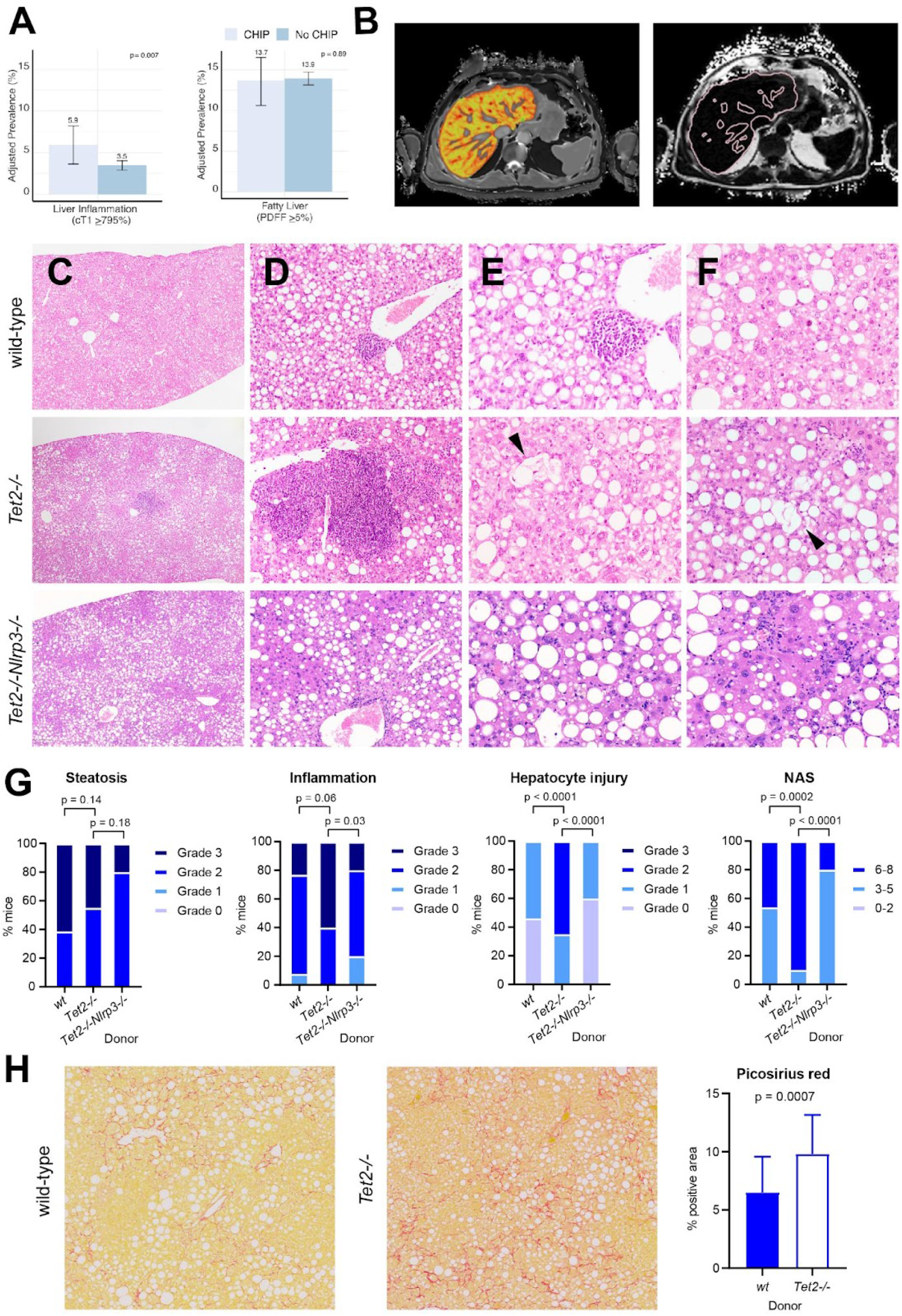
Clonal hematopoiesis of indeterminate potential is associated with steatohepatitis. **(A)** Prevalence of liver inflammation and hepatic steatosis on magnetic resonance imaging among 8251 individuals in the UK Biobank. Liver inflammation was defined as a corrected T1 signal ≥ 795 ms. Fatty liver was defined as a proton density fat fraction ≥ 5%. Logistic regression, with adjustment for age and sex, was used to test the association between CHIP status and the presence of fatty liver and liver inflammation. **(B)** Perspectum Multiscan corrected T1 image (left) and proton density fat fraction (right) of a patient with CHIP. Reproduced by kind permission of the UK Biobank. **(C-G)** B6.SJL mice were transplanted with *Tet2*^*-/-*^ (n=20), *Tet2*^*-/-*^*Nlrp3*^*-/-*^ (n=10) or control *vavCre*^*+*^ (n=13) bone marrow cells and fed CDAHFD for 11 weeks. Steatohepatitis was graded using modified NASH CRN histologic criteria. Compared to *vavCre*^*+*^ and *Tet2*^*-/-*^*Nlrp3*^*-/-*^ animals, *Tet2*^*-/-*^ transplanted mice show similar accumulation of liver fat (**C**) and show increased lymphocyte and macrophage inflammation (**D**) and increased hepatocyte ballooning injury (**E-F**, arrowheads), resulting in higher aggregate NAFLD activity score (NAS) (**G**). **(H)** B6.SJL mice were transplanted with *Tet2*^*-/-*^ (n=24) or *vavCre*^*+*^ wild type control (n=21) bone marrow cells and fed CDAHFD for 19 weeks. Collagen fibrosis, highlighted by Picosirius red staining, was quantified as the percent positive area using ImageJ. CHIP, clonal hematopoiesis of indeterminate potential; cT1, corrected T1; NAS, nonalcoholic fatty liver disease activity score; PDFF, proton density fat fraction.

### Mendelian randomization analysis of CHIP association with chronic liver disease

To assess whether the association of CHIP status with chronic liver disease is causal, we performed Mendelian randomization analysis. We identified 184 independent genetic variants associated with CHIP status at p<0.0001 significance from a recent GWAS comprised of 97,691 blood DNA-derived whole genome sequences.(*20*) We tested the association of these variants with cirrhosis risk using summary statistics from a genome wide association study of 5770 cirrhosis cases and 487780 controls.(*17*) Using the MR-RAPS method, which increases power to detect a significant effect of genetic predisposition to CHIP and accounts for potential directional pleiotropy, CHIP was associated with a three-fold risk of chronic liver disease (OR 3.20 CI 1.95, 5.24, p<0.001). This estimate did not differ significantly from the observational estimate (p_interaction_=0.78, **Supp. Figure 9**). Similar estimates were obtained in sensitivity analyses using the pleiotropy-robust Mendelian randomization methods median regression and MR-PRESSO, multivariate MR, as well as using MR-RAPS with varying p-value thresholds for SNP instrument inclusion (**Supp. Table 5**). As confirmation of the robustness of the MR-RAPS framework, genetic predisposition to CHIP was significantly associated with the development of myeloproliferative neoplasms (OR 39.4 CI 5.2, 188.0, p<0.001), consistent with the known association between CHIP and hematologic malignancies.*(10, 33)*

### Effect of Tet2^-/-^ hematopoiesis on diet induced steatohepatitis in mice

To examine the role of *Tet2* deficiency in causing chronic liver inflammation, we utilized mouse models of steatohepatitis in which fatty liver and chronic inflammation are induced by diet. Choline deficient, amino acid defined, high fat diet (CDAHFD) disrupts hepatic mitochondrial beta oxidation and production of very low density lipoprotein, resulting in liver oxidative damage and chronic liver injury.(*23, 24*) B6.SJL mice were transplanted with *Tet2*^*-/-*^ or control bone marrow cells and fed CDAHFD for 11 weeks, after which liver fat, inflammation and hepatocyte ballooning injury were assessed histologically and integrated into a modified nonalcoholic fatty liver disease activity score (NAS) (**Supp. Table 7**). Mice with wild-type or*Tet2*^*-/-*^ hematopoietic cells had similar hematologic parameters and liver fat accumulation (**Figure 2C, Supp. Figure 10**). In contrast, *Tet2*^-/-^ transplanted animals showed more lobular inflammation with prominent lymphoid aggregates and more hepatocyte ballooning injury (**Figure 2D-F**), corresponding to overall higher NAS (**Figure 2G**). These findings demonstrate that *Tet2*^-/-^ hematopoietic cells promote the development of steatohepatitis in mice. Similar findings of increased liver inflammation and increased hepatocyte damage were observed in hypercholesterolemic *Ldlr*^-/-^ mice transplanted with *Tet2*^-/-^ bone marrow cells and fed Western diet (**Supp. Figure 11A-F**), indicating that the effect of hematopoietic *Tet2* loss is consistent across different dietary and genetic models that promote steatohepatitis. Furthermore, transplantation of hematopoietic cells lacking *Dnmt3a*, the most commonly mutated gene in CHIP, similarly led to increased liver inflammation and higher aggregate NAS in CDAHFD fed mice (**Supp. Figure 11G**). Therefore, we reproducibly found that mutant hematopoietic cells lacking *Tet2* or *Dnmt3a* aggravate chronic liver injury in models of steatohepatitis.

To study the persistence of liver injury in the setting of mutant hematopoiesis, *Tet2*^-/-^ transplanted animals were fed CDAHFD for 11 weeks and subsequently fed regular chow for 10 days. We observed a global decrease in steatohepatitis after diet reversion, as reflected in lower NAS scores; nevertheless, liver inflammation and hepatocyte injury persisted at significant levels in *Tet2*^-/-^ transplanted mice compared to wild type controls (**Supp. Figure 11H**). In contrast, liver fat regressed at a similar rate to control mice. These findings suggest that the prolongation of liver inflammation and injury by *Tet2*^-/-^ hematopoietic cells may promote chronic liver disease in the setting of repeated liver insults.(*25*) Persistent liver inflammation can also stimulate inflammatory scarring and fibrosis of the liver.(*4*) Liver fibrosis was not observed in *Tet2*^-/-^ and control animals fed CDAHFD for 11 weeks (**Supp. Figure 11F**); therefore, mice were diet fed for an extended duration. At 19 weeks, *Tet2*^-/-^ transplanted mice showed significantly increased liver fibrosis compared to wild type controls (**Figure 2H**). This observation confirmed that hematopoietic loss of *Tet2* promotes the progression of steatohepatitis to liver fibrosis in CDAHFD fed mice.

### CHIP mediates chronic liver injury via proinflammatory signaling

Prior work revealed that a damaging interleukin 6 receptor gene (*IL6R)* missense variant (p.Asp358Ala, rs2228145) is associated with greater protection from coronary artery disease among individuals with CHIP versus individuals without CHIP.(*19*) Therefore, we examined whether this variant is also associated with protection from chronic liver disease among individuals with CHIP. We observed a significant interaction in the association of Asp358Ala with chronic liver disease by CHIP status (p_interaction_=0.03). Asp358Ala was not associated with chronic liver disease risk among those without CHIP (OR 1.17 CI 0.91, 1.50, p=0.21). In contrast, Asp358Ala protected against chronic liver disease among individuals with CHIP (OR 0.32 CI 0.11, 0.95, p=0.04) (**Figure 3A**). This finding implicates proinflammatory IL6 signaling in the contribution of CHIP to chronic liver disease.

**Figure 3.**
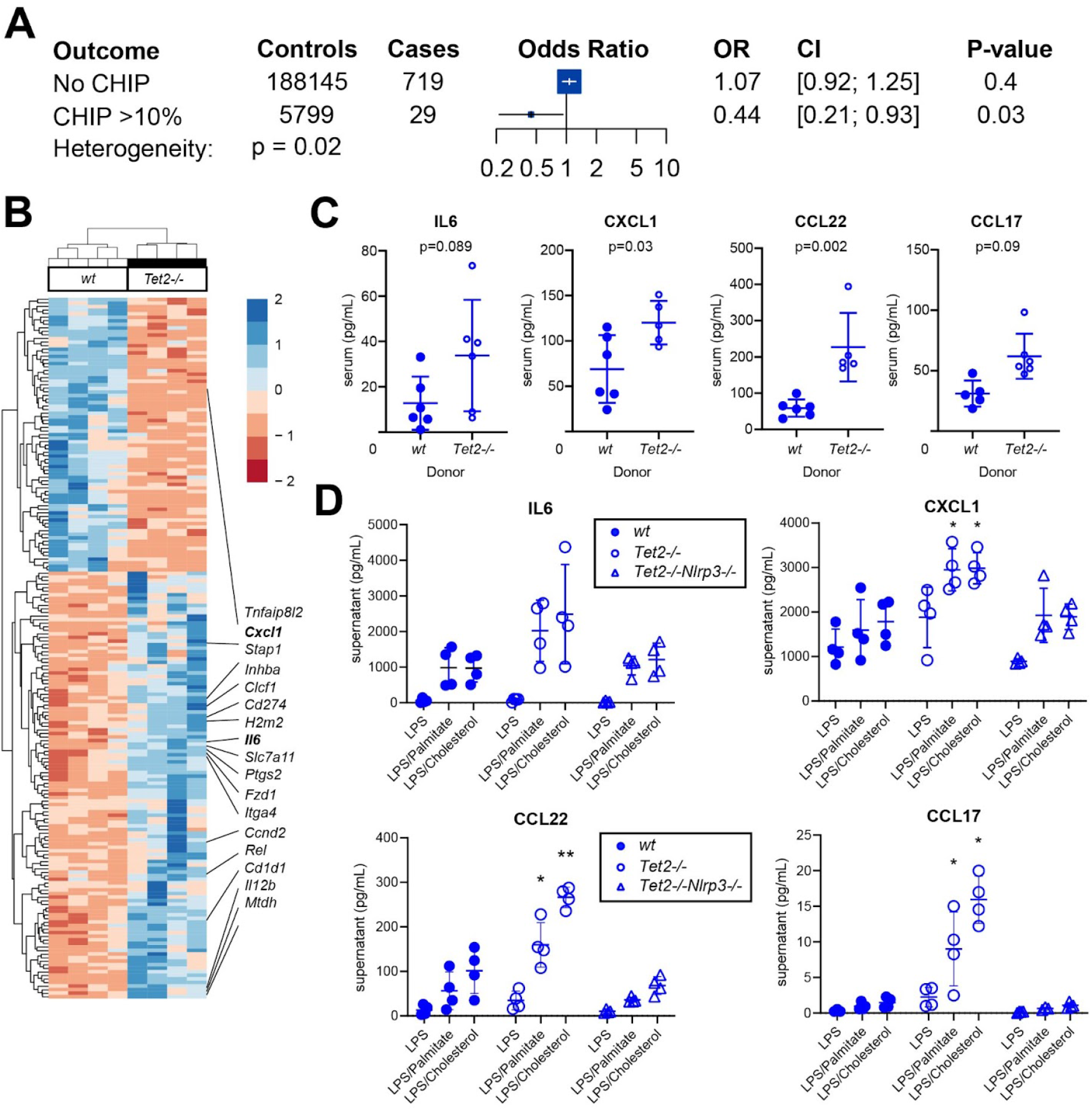
Proinflammatory signaling in clonal hematopoiesis of indeterminate potential. **(A)** Association of *IL6R* p.Asp358Ala germline mutation with chronic liver disease in CHIP (variant allele fraction ≥ 10%) versus individuals without CHIP. **(B)** Unsupervised hierarchical clustering of differentially regulated genes in sorted liver macrophages from B6.SJL mice transplanted with *Tet2*^*-/-*^ (n=4) or control *vavCre*^*+*^ (*wt*; n=4) bone marrow cells and fed CDAHFD for 11 weeks. **(C)** After 19 weeks of CDAHFD, *Tet2*^*-/-*^ or control (*wt*) transplanted mice were bled and serum obtained for cytokine measurements. **(D)** Bone marrow derived macrophages (BMDM) from *Tet2*^*-/-*^, *Tet2*^*-/-*^*Nlrp3*^*-/-*^ or control *vavCre*^*+*^ (*wt*) mice were primed with low dose lipopolysaccharide (LPS) for 2 hours and stimulated with palmitic acid or cholesterol monohydrate crystals as indicated for 6 hours. Cytokine levels in tissue culture supernatant were measured and showed increased expression of IL6, CXCL1, CCL22 and CCL17 in *Tet2*^*-/-*^ transplanted, but not *Tet2*^*-/-*^*Nlrp3*^*-/-*^ transplanted mice. CHIP, clonal hematopoiesis of indeterminate potential; CI, 95% confidence interval; LPS, lipopolysaccharide; OR, odds ratio.

IL6 is a potent inflammatory cytokine under the transcriptional control of IL-1β and mediates numerous immune responses downstream of the NLRP3 inflammasome.(*26*) Therefore, we tested whether proinflammatory signaling via NLRP3 contributes to the effect of hematopoietic *Tet2* loss. Compared to mice transplanted with *Tet2*^-/-^ bone marrow, mice transplanted with *Tet2*^-/-^*Nlrp3*^-/-^ bone marrow and fed CDAHFD for 11 weeks showed significantly reduced liver inflammation and hepatocyte injury, resulting in overall lower NAS comparable to control transplanted mice (**Figure 2B-E**). This result demonstrates that *Tet2*^*-/-*^ dependent steatohepatitis requires NLRP3 inflammasome.

### Tet2 dependent inflammatory signaling in monocyte-derived liver macrophages

Liver resident phagocytes, namely Kupffer cells, are a major source of proinflammatory cytokine secretion in response to immune stimuli and inflammasome activation. After hematopoietic transplant, Kupffer cells, which express CD45^+^F4/80^+^CD11b^int^, are replaced by monocyte derived CD45.2 macrophages, which express CD45^+^F4/80^mod^CD11b^hi^ (**Supp. Figure 12A**).(*27*)(*28*) We performed RNA sequencing on CD45^+^F4/80^mod^CD11b^hi^ monocyte derived liver macrophages in CDAHFD-fed mice transplanted with *Tet2*^-/-^ or control bone marrow (**Supp.Table 7-8**). Compared to wild type cells, *Tet2*^-/-^ macrophages showed increased expression of *Il6* and *Cxcl1* among other pro-inflammatory genes (**Figure 3B**). Consistent with transcriptional analysis, mice transplanted with *Tet2*^-/-^ hematopoietic cells showed increased serum levels of IL6, CXCL1, CCL22 and CCL17 (**Figure 3C**). CXCL1 and IL6 are pro-inflammatory molecules regulated by the NLRP3 inflammasome complex, whereas the related chemokines CCL17 and CCL22 signal through CCR4 to promote the recruitment of regulatory T cells.(*29*) Confirming the key role of NLRP3 in mediating downstream proinflammatory cytokine secretion, bone marrow derived macrophages lacking both *Tet2* and *Nlrp3* showed baseline expression levels of IL6, CXCL1, CCL22 and CCL17 (**Figure 3D**). Therefore, hematopoietic *Tet2* loss exerts its proinflammatory effect via downstream cytokine secretion in an NLRP3 dependent fashion. Gene set enrichment analysis further confirmed that *Tet2*^-/-^ liver macrophages exhibit pro-inflammatory activity via increased IL6 and TNFα signaling, as well as identifying potential pathways to fibrosis via enhanced TGFβ and WNT signaling (**Supp. Figure 12B**). In addition, bulk liver mRNA from mice transplanted with *Tet2*^-/-^ marrow show enrichment in transcriptional programs associated with steatohepatitis and liver fibrosis (**Supp. Figure 12C, Supp. Table 10-11**). Together, these findings support a model in which *Tet2* deficient hematopoiesis induces NLRP3 dependent proinflammatory signals in liver resident immune cells to promote steatohepatitis and fibrosis.

## Discussion

By combining large-scale human genetic studies with *in vivo* modeling of clonal hematopoiesis in murine models, we demonstrate that CHIP is associated with an elevated risk of chronic liver disease, including NASH, via aberrant inflammatory responses. Among middle-aged adults without liver disease in the UK Biobank, 1 in 20 individuals with CHIP developed chronic liver disease by 80 years of age, compared to only 1 in 100 individuals without CHIP. The overall nearly four-fold risk of incident chronic liver disease observed in the current study is greater than the nearly two-fold risk of incident coronary artery disease previously reported with CHIP in overlapping cohorts.(*12, 19*) Moreover, it is generally accepted that medical billing codes incompletely capture chronic liver disease in patients, suggesting the absolute increase in chronic liver disease with CHIP may be underestimated. Mendelian randomization and mouse studies support a causal role for CHIP in the pathogenesis of chronic liver disease. Furthermore, Mendelian randomization analyses and *in vivo* inflammatory biomarker and transcriptional analyses implicate NLRP3 inflammasome and downstream IL-6 activity in CHIP-associated chronic liver disease.

Our findings support a model of CHIP promoting steatohepatitis particularly among individuals with elevated liver fat or other sources of liver injury that increase the risk of cirrhosis.(*25*) First, individuals with CHIP showed higher indices of liver inflammation and fibrosis with no difference in liver fat accumulation. Second, hematopoietic-specific *Tet2* inactivation increased the severity of diet-induced steatohepatitis in mice due to increased liver inflammation, hepatocyte injury and fibrosis, without influence on liver fat. In contrast to germline genetic variants which predispose to both liver fat and cirrhosis,(*17*) these findings are compatible with the tendency of clonal hematopoiesis to exaggerate the proinflammatory response to an existing stimulus, which in the case of NASH results in the activation of local immune and fibrogenic pathways in the fatty liver. Nevertheless, liver fat accumulation may elicit different inflammatory responses in normal and clonal hematopoiesis.

We also provide human genetic evidence for a causal relationship between CHIP and liver disease. Mendelian randomization analysis may be useful to distinguish outcomes that are caused by CHIP from other confounding associations. To optimize statistical power in the setting of low heritability of CHIP(*20*), we applied a newly developed Mendelian randomization technique (MR-RAPS), which allows for the use of sub-genome wide significant variants.(*21*) We observed that germline genetic predisposition to CHIP predisposes to chronic liver disease risk. Therefore, CHIP is likely a causal risk factor for chronic liver disease. Consequently, targeting factors that promote CHIP-associated liver injury as well as targeting CHIP itself are both expected to reduce the risk of chronic liver disease among susceptible individuals.

Genetic deficiency of IL-6 signaling due to the presence of *IL6R* p.Asp358Ala in CHIP individuals was associated with a greater reduction in chronic liver disease risk, but not among those without CHIP. This finding is compatible with our prior observation that the presence of *IL6R* p.Asp358Ala was associated with a markedly reduced risk for incident cardiovascular disease risk, but not hematologic malignancy, specifically among individuals with CHIP.(*19*) In the current study, we observed an insignificant trend towards increased serum IL6 levels in *Tet2*^-/-^ bone marrow transplanted mice, and *Tet2*^-/-^ liver macrophages also exhibited higher levels of IL6 expression in addition to other proinflammatory cytokines and chemokines with proven roles in chronic liver disease pathogenesis. The expression of these proinflammatory cytokines required the upstream regulator NLRP3, and the development of steatohepatitis in *Tet2*^*-/-*^ transplanted mice was abrogated by the genetic loss of NLRP3. Although the role of IL6 in NAFLD and other chronic liver diseases remains controversial,(*30, 31*) broad pharmacologic inhibition of NLRP3 inflammasome reduces liver inflammation and fibrosis in diet-fed mice.(*32*) These findings suggest that therapies targeting inflammatory pathways may be useful for prevention of chronic liver disease especially in individuals with CHIP.

In conclusion, CHIP is associated with an elevated risk of chronic liver disease specifically through the promotion of liver inflammation and injury. Through targeting of the NLRP3 inflammasome or downstream mediators, CHIP may be a modifiable risk factor for chronic liver disease.

## Data Availability

All data produced in the present work are contained in the manuscript.

## Acknowledgements

The UK Biobank analyses were performed under application #7089 and #50834. The investigators would like to thank the UK Biobank staff and participants. P.N. is supported by a Hassenfeld Scholar Award and the Paul & Phyllis Fireman Endowed Chair in Vascular Medicine from the Massachusetts General Hospital, and grants from the National Heart, Lung, and Blood Institute (R01HL142711, R01HL148565, and R01HL148050) and National Institute of Diabetes and Digestive and Kidney Diseases (R01DK125782). P.N. and B.L.E. are supported by a grant from the Fondation Leducq (TNE-18CVD04). B.L.E was also supported by the NIH (R01HL082945, P01CA108631, and P50CA206963) and the Howard Hughes Medical Institute. S.M.Z is supported by the NIH National Heart, Lung, and Blood Institute (1F30HL149180-01) and the NIH Medical Scientist Training Program Training Grant (T32GM136651). R.S.S. is supported by a Kay Kendall Leukaemia Fund Intermediate Fellowship and by a CRUK Advanced Clinician Scientist Fellowship. M.A. is supported by the Deutsche Forschungsgemeinschaft (DFG, AG252/1-1).

Molecular data for the Trans-Omics in Precision Medicine (TOPMed) program was supported by the National Heart, Lung and Blood Institute (NHLBI). See the TOPMed Omics Support Table for study specific omics support information. Core support including centralized genomic read mapping and genotype calling, along with variant quality metrics and filtering were provided by the TOPMed Informatics Research Center (3R01HL-117626-02S1; contract HHSN268201800002I). Core support including phenotype harmonization, data management, sample-identity QC, and general program coordination were provided by the TOPMed Data Coordinating Center (R01HL-120393; U01HL-120393; contract HHSN268201800001I). We gratefully acknowledge the studies and participants who provided biological samples and data for TOPMed.

## TOPMed Omics Support Table

| TOPMed Accession # | TOPMed Project | Parent Study | TOPMed Phase | Omics Center | Omics Support | Omics Type |
| --- | --- | --- | --- | --- | --- | --- |
| phs000974 | AFGen | FHS<br>AFGen | 1 | Broad Genomics | 3R01HL092577-06S1 | WGS |
| phs000974 | FHS | FHS | pilot; 4.5 | NWGC | HHSN268201600032I | RNASeq |
| phs000974 | FHS | FHS | 5 | NWGC | HHSN268201600032I | RNASeq |
| phs000974 | FHS | FHS | pilot; 4.5 | Broad BIDMC Proteomics | HHSN268201600034I | Proteomics |
| phs000974 | FHS | FHS | 5 | Broad Metabolomics | HHSN268201600034I | Metabolomics |
| phs000974 | FHS | FHS | 5 | Keck MGC | HHSN268201600038I | Methylomics |
| phs000974 | FHS | FHS | 1 | Broad Genomics | 3U54HG003067-12S2 | WGS |

We thank Dr. Darrick K. Li and Dr. Peter G. Miller for experimental advice and critical reading of the manuscript.

## Author Contributions

W.W., C.E., B.L.E., and P.N. conceived of, designed, and wrote the manuscript for this project. B.L.E. and P.N. provided supervision, project administration, and secured funding. C.E. led human genetics data curation and analyses, and W.W. led animal experiments and analyses. A.B., S.Z., C.G., J.P., M.U., A.N. contributed to human genetic data and revised the manuscript for intellectual content. All other authors revised the manuscript for intellectual content.

