## Supplementary materials for "Clonal hematopoiesis and risk of chronic liver disease"

**Supplementary Appendix**

**Supplementary Tables**

**Supplementary Table 1.** Chronic liver diseases cases and definitions in five samples.
**Supplementary Table 2.** CHIP variants ascertained through whole-exome sequencing.
**Supplementary Table 3.** CHIP variants ascertained through genotyping.
**Supplementary Table 4.** Association of CHIP with known risk factors across study cohorts.
**Supplementary Table 5.** Mendelian randomization analyses of CHIP associations with chronic liver disease.
**Supplementary Table 6.** Genetic variants used for Mendelian randomization analyses.
**Supplementary Table 7.** Histologic grading criteria for steatohepatitis in mice.
**Supplementary Table 8.** Differentially regulated genes in sorted liver macrophages from *Tet2*$^{-/-}$ and control bone marrow transplanted mice
**Supplementary Table 9.** Gene set enrichment analysis of *Tet2*$^{-/-}$ versus control sorted liver macrophages.
**Supplementary Table 10.** Differentially regulated genes in bulk liver transcripts from *Tet2*$^{-/-}$ and control bone marrow transplanted mice.
**Supplementary Table 11.** Gene set enrichment analysis of *Tet2*$^{-/-}$ versus control bulk liver RNA.

**Supplementary Figures**

**Supplementary Figure 1.** Study design of chronic liver disease associations with CHIP.
**Supplementary Figure 2**. Proportion of CHIP by mutated gene among 3068 individuals with CHIP.
**Supplementary Figure 3.** Prevalence of CHIP by age.
**Supplementary Figure 4.** Association of CHIP with prevalent chronic liver disease by variant allele fraction.
**Supplementary Figure 5.** Association of CHIP with prevalent chronic liver disease by mutated gene.
**Supplementary Figure 6.** Cumulative risk of chronic liver disease by clonal hematopoiesis status in the UK Biobank.
**Supplementary Figure 7.** Association of CHIP with serum biomarkers in the UK Biobank.
**Supplementary Figure 8.** Association of CHIP with serum biomarkers in the UK Biobank with carriers of *JAK2*-mutant CHIP excluded.
**Supplementary Figure 9.** Mendelian randomization analysis of CHIP association with chronic liver disease.
**Supplementary Figure 10.** Liver and hematologic parameters of *Ldlr-/-* (A-F) and B6.SJL (G-L) mice transplanted with *Tet2-/-* or control bone marrow cells and fed Western diet for 9 weeks or CDAHFD for 10 weeks, respectively.

**Supplementary Figure 11.** Steatohepatitis and liver fibrosis in *Tet2*$^{-/-}$ and *Dnmt3a*$^{-/-}$ transplanted mice.
**Supplementary Figure 12.** Gene expression analysis of sorted liver macrophages and bulk liver RNA in *Tet2*$^{-/-}$ transplanted mice fed CDAHFD.

**Extended Materials and Methods**

**References**

**Supplementary Table 1. Chronic liver diseases cases and definitions in five samples**.

| Cohort | Cases | Controls | Definition | Analysis |
|---|---|---|---|---|
| Framingham Heart Study | 46 | 4194 | Prevalent cirrhosis or chronic liver disease, physician-diagnosed or ICD codes K70.2, K70.3, K70.4, K74.0, K74.1, K74.2, K74.6, K76.6, or I85 | Observational |
| Atherosclerosis Risk in Communities Study | 33 | 7381 | Prevalent cirrhosis or chronic liver disease, physician-diagnosed or ICD codes K70.2, K70.3, K70.4, K74.0, K74.1, K74.2, K74.6, K76.6, or I85 | Observational |
| UK Biobank WES | 555 | 200,854 | Prevalent chronic liver disease, hospitalization or death due to ICD codes K70.2, K70.3, K70.4, K74.0, K74.1, K74.2, K74.6, K76.6, or I85 | Observational |
| UK Biobank Array | 825 | 238,491 | Incident chronic liver disease, hospitalization or death due to ICD codes K70.2, K70.3, K70.4, K74.0, K74.1, K74.2, K74.6, K76.6, or I85 | Observational |
| MGB Biobank | 114 | 1386 | Biopsy-diagnosed non-alcoholic steatohepatitis | Observational |
| Multi-cohort GWAS (*Emdin et al.*) | 5777 | 487780 | Prevalent chronic liver disease, hospitalization or death due to cirrhosis | Mendelian randomization |

GWAS, genome-wide association study; WES, whole exome sequencing

**Supplementary Table 2. CHIP variants ascertained through whole-exome sequencing.**

| Gene name | Reported mutations used for variant calling | Accession |
|---|---|---|
| *ASXL1* | Frameshift/nonsense/splice-site in exon 11-12 | NM_015338 |
| *ASXL2* | Frameshift/nonsense/splice-site in exon 11-12 | NM_018263 |
| *BCOR* | Frameshift/nonsense/splice-site | NM_001123385 |
| *BCORL1* | Frameshift/nonsense/splice-site | NM_021946 |
| *BRAF* | G464E, G464V, G466E, G466V, G469R, G469E, G469A, G469V, V471F, V472S, L485W, N581S, I582M, I592M, I592V, D594N, D594G, D594V, D594E, F595L, F595S, G596R, L597V, L597S, L597Q, L597R, A598V, V600M, V600L, V600K, V600R, V600E, V600A, V600G, V600D, K601E, K601N, R603*, W604R, W604G, S605G, S605F, S605N, G606E, G606A, G606V, H608R, H608L, G615R, S616P, S616F, L618S, L618W | NM_004333 |
| *BRCC3* | Frameshift/nonsense/splice-site | NM_024332 |
| *CBL* | RING finger missense p.381-421 | NM_005188 |
| *CBLB* | RING finger missense p.372-412 | NM_170662 |
| *CEBPA* | Frameshift/nonsense/splice-site | NM_004364 |
| *CREBBP* | Frameshift/nonsense/splice-site, D1435E, R1446L, R1446H, R1446C, Y1450C, P1476R, Y1482H, H1487Y, W1502C, Y1503D, Y1503H, Y1503F, S1680del | NM_004380 |
| *CSF1R* | L301F, L301S, Y969C, Y969N, Y969F, Y969H, Y969D | NM_005211 |
| *CSF3R* | T615A, T618I, truncating c.741-791 | NM_000760 |
| *CTCF* | Frameshift/nonsense, R377C, R377H, P378A, P378L | NM_006565 |
| *CUX1* | Frameshift/nonsense | NM_181552 |
| *DNMT3A* | Frameshift/nonsense/splice-site, F290I, F290C, V296M, P307S, P307R, R326H, R326L, R326C, R326S, G332R, G332E, V339A, V339M, V339G, L344Q, L344P, R366P, R366H, R366G, A368T, A368V, R379H, R379C, I407T, I407N, I407S, F414L, F414S, F414C, A462V, K468R, C497G, C497Y, Q527H, Q527P, Y533C, S535F, C537G, | NM_022552 |

| | | |
|---|---|---|
| | C537R, G543A, G543S, G543C, L547H, L547P, L547F, M548I, M548K, G550R, W581R, W581G, W581C, R604Q, R604W, R635W, R635Q, S638F, G646V, G646E, L653W, L653F, I655N, V657A, V657M, R659H, Y660C, V665G, V665L, M674V, R676W, R676Q, G685R, G685E, G685A, D686Y, D686G, R688H, G699R, G699S, G699D, P700L, P700S, P700R, P700Q, P700T, P700A, D702N, D702Y, V704M, V704G, I705F, I705T, I705S, I705N, G707D, G707V, C710S, C710Y, S714C, V716D, V716F, V716I, N717S, N717I, P718L, R720H, R720G, K721R, K721T, Y724C, R729Q, R729W, R729G, F731C, F731L, F731Y, F731I, F732del, F732C, F732S, F732L, E733G, E733A, F734L, F734C, Y735C, Y735N, Y735S, R736H, R736C, R736P, L737H, L737V, L737F, L737R, A741V, P742P, P743R, P743L, R749C, R749L, R749H, R749G, F751L, F751C, F752del, F752C, F752L, F752I, F752V, W753G, W753C, W753R, L754P, L754R, L754H, F755S, F755I, F755L, M761I, M761V, G762C, V763I, S770L, S770W, S770P, R771Q, F772I, F772V, L773R, L773V, E774K, E774D, E774G, I780T, D781G, R792H, W795C, W795L, G796D, G796V, N797Y, N797H, N797S, P799S, P799R, P799H, R803S, R803W, P804L, P804S, K826R, S828N, K829R, T835M, N838D, K841Q, Q842E, P849L, D857N, W860R, E863D, F868S, G869S, G869V, M880V, S881R, S881I, R882H, R882P, R882C, R882G, A884P, A884V, Q886R, L889P, L889R, G890D, G890R, G890S, V895M, P896L, V897G, V897D, R899L, R899H, R899C, L901R, L901H, P904L, F909C, P904Q, A910P, C911R, C911Y | |
| *EED* | Frameshift/nonsense/splice-site, L240Q, I363M | NM_003797 |
| *EP300* | Frameshift/nonsense/splice_site, VF1148_1149del, D1399N, D1399Y, P1452L, Y1467N, Y1467H, Y1467C, R1627W, A1629V | NM_001429 |
| *ETNK1* | N244S, N244T, N244K | NM_018638 |
| *ETV6* | Frameshift/nonsense/splice-site | NM_001987 |
| *EZH2* | Frameshift/nonsense/splice-site, Q62R, N102S, F145S, F145C, F145Y, F145L, G159R, E164D, R202Q, K238E, E244K, R283Q, H292R, P488S, R497Q, R561H, T568I, K629E, Y641N, Y641H, Y641S, Y641C, Y641F, D659Y, | NM_001203247 |

| | | |
|---|---|---|
| | D659G, V674M, A677G, A677V, R679C, R679H, R685C, R685H, A687V, N688I, N688K, H689Y, S690P, I708V, I708T, I708M, E720K, E740K | |
| *FLT3* | V579A, V592A, V592I, F594L, FY590-591GD, D835Y, D835H, D835E, del835 | NM_004119 |
| *GATA1* | Frameshift/nonsense/splice-site | NM_002049 |
| *GATA2* | Frameshift/nonsense/splice-site, R293Q, N317H, A318T, A318V, A318G, G320D, L321P, L321F, L321V, Q328P, R330Q, R361L, L359V, A372T, R384G, R384K | NM_001145661 |
| *GATA3* | Frameshift/nonsense/splice-site ZNF domain, R276W, R276Q, N286T, L348V, | NM_001002295 |
| *GNA13* | I34T, G57S, S62F, M68K, Q134R, Y145F, L152F, E167D, Q169H, R264H, E273K, V322G, V362G, L371F | NM_006572 |
| *GNAS* | R201S, R201C, R201H, R201L, Q227K, Q227R, Q227L, Q227H, R374C | NM_000516 |
| *GNB1* | K57N, K57M, K57E, K57T, I80T, I80N | NM_002074 |
| *IDH1* | R132C, R132G, R132H, R132L, R132P, R132V, V178I | NM_005896 |
| *IDH2* | R140W, R140Q, R140L, R140G, R172W, R172G, R172K, R172T, R172M, R172N, R172S | NM_002168 |
| *IKZF1* | Frameshift/nonsense | NM_006060 |
| *IKZF2* | Frameshift/nonsense | NM_016260 |
| *IKZF3* | Frameshift/nonsense | NM_012481 |
| *JAK1* | T478A, T478S, V623A, A634D, L653F, R724H, R724Q, R724P, T782M, L783F | NM_002227 |
| *JAK2* | N533D, N533Y, N533S, H538R, K539E, K539L, I540T, I540V, V617F, R683S, R683G, del/ins537-539L, del/ins538-539L, del/ins540-543MK, del/ins540-544MK, del/ins541-543K, del542-543, del543-544, ins11546-547 | NM_004972 |
| *JAK3* | M511T, M511I, A572V, A572T, A573V, R657Q, V715I, V715A | NM_000215 |

| | | |
|---|---|---|
| *KDM6A* | Frameshift/nonsense/splice-site, del419 | NM_021140 |
| *KIT* | ins503, V559A, V559D, V559G, V559I, V560D, V560A, V560G, V560E, del560, E561K, del579, P627L, P627T, R634W, K642E, K642Q, V654A, V654E, H697Y, H697D, E761D, K807R, D816H, D816Y, D816F, D816I, D816V, D816H, del551-559 | NM_000222 |
| *KRAS* | G12D, G12A, G12E, G12V, G13D, G13C, G13Y, G13F, G13R, G13A, G13V, G13E, V14I, T58I, G60D, G60A, G60V, Q61K, Q61E, Q61P, Q61R, Q61L, Q61H, K117E, K117N, A146T, A146P, A146V | NM_033360 |
| *LUC7L2* | Frameshift/nonsense/splice-site | NM_016019 |
| *MLL* | Frameshift/nonsense | NM_005933 |
| *MLL2* | Frameshift/nonsense | NM_003482 |
| *MPL* | S505G, S505N, S505C, L510P, del513, W515A, W515R, W515K, W515S, W515L, A519T, A519V, Y591D, W515-518KT | NM_005373 |
| *NF1* | Frameshift/nonsense | NM_000267 |
| *NPM1* | Frameshift p.W288fs (insertion at c.859_860, 860_861, 862_863, 863_864) | NM_002520 |
| *NRAS* | G12S, G12R, G12C, G12N, G12P, G12Y, G12D, G12A, G12V, G12E, G13S, G13R, G13C, G13N, G13P, G13Y, G13D, G13A, G13V, G13E, G60E, G60R, Q61R, Q61L, Q61K, Q61P, Q61H, Q61Q | NM_002524 |
| *PDS5B* | Frameshift/nonsense/splice-site, R1292Q | NM_015032 |
| *PDSS2* | Frameshift/nonsense | NM_020381 |
| *PHF6* | Frameshift/nonsense/splice-site, A40D, M125I, S246Y, F263L, R274Q, C297Y, H302Y, H329L | NM_001015877 |
| *PHIP* | Frameshift/nonsense/splice-site | NM_017934 |
| *PPM1D* | Frameshift/nonsense, exon 5 or 6 | NM_003620 |
| *PRPF40B* | Frameshift/nonsense/splice-site, P15H, M58I, P405L, P562S, | NM_001031698 |

| | | |
|---|---|---|
| *PRPF8* | M1307I, C1594W, D1598Y, D1598N, D1598V | NM_006445 |
| *PTEN* | Frameshift/nonsense/splice-site, D24G, R47G, F56V, L57W, H61R, K66N, Y68H, C71Y, F81C, Y88C, D92G, D92V, D92E, H93Y, H93D, H93Q, N94I, P95L, I101T, C105F, C105S, D107Y, L112V, H123Y, C124R, C124S, K125E, A126D, K128N, R130G, R130Q, R130L, G132D, I135V, I135K, C136R, C136F, K144Q, A151T, D153Y, D153N, Y155H, Y155C, R159K, R159S, R161K, R161I, G165R, G165E, S170N, S170I, R173C, Y174D, Y177C, H196Y, R234W, G251C, D252Y, F271S, D326G | NM_000314 |
| *PTPN11* | G60V, G60R, G60A, D61Y, D61V, D61G, Y63C, E69K, E69G, E69D, E69Q, F71L, F71K, A72T, A72V, A72D, T73I, E76K, E76Q, E76M, E76A, E76G, E139G, E139D, N308D, N308T, N339S, P491L, S502P, S502A, S502L, G503V, G503G, G503A, G503E, Q506P, T507A, T507K | NM_002834 |
| *RAD21* | Frameshift/nonsense/splice-site, R65Q, H208R, Q474R | NM_006265 |
| *RUNX1* | Frameshift/nonsense/splice-site, S73F, H78Q, H78L, R80C, R80P, R80H, L85Q, P86L, P86H, S114L, D133Y, L134P, R135G, R135K, R135S, R139Q, R142S, A165V, R174Q, R177L, R177Q, A224T, D171G, D171V, D171N, R205W, R223C | NM_001001890 |
| *SETBP1* | D868N, D868T, S869N, G870S, I871T, D880N, D880Q | NM_015559 |
| *SETD2* | Frameshift/nonsense, V1190M | NM_014159 |
| *SETDB1* | Frameshift/nonsense, K715E | NM_001145415 |
| *SF1* | Frameshift/nonsense/splice-site, T454M, Y476C, A508G | NM_004630 |
| *SF3A1* | Frameshift/nonsense/splice-site, A57S, M117I, K166T, Y271C | NM_005877 |
| *SF3B1* | G347V, R387W, R387Q, E592K, E622D, Y623C, R625L, R625C, R625G, H662Q, H662D, T663I, K666N, K666T, K666E, K666R, K700E, V701F, A708T, G740R, G740E, A744P, D781G, E783K, R831Q, L833F, E862K, R957Q | NM_012433 |
| *SRSF2* | Y44H, P95H, P95L, P95T, P95R, P95A, P107H, P95fs | NM_003016 |
| *SMC1A* | K190T, R586W, M689V, R807H, R1090H, R1090C | NM_006306 |

| | | |
|---|---|---|
| *SMC3* | Frameshift/nonsense, R155I, Q367E, D392V, K571R, R661P, G662C | NM_005445 |
| *STAG1* | Frameshift/nonsense/splice-site, H1085Y | NM_005862 |
| *STAG2* | Frameshift/nonsense/splice-site | NM_006603 |
| *SUZ12* | Frameshift/nonsense | NM_015355 |
| *TET2* | Frameshift/nonsense/splice-site, missense mutations in catalytic domains (p.1104-1481 and 1843-2002) | NM_001127208 |
| *TP53* | Frameshift/nonsense/splice-site, S46F, G105C, G105R, G105D, G108S, G108C, R110L, R110C, T118A, T118R, T118I, S127F, S127Y, L130V, L130F, K132Q, K132E, K132W, K132R, K132M, K132N, F134V. F134L, F134S, C135W, C135S, C135F, C135G, C135Y, Q136K, Q136E, Q136P, Q136R, Q136L, Q136H, A138P, A138V, A138A, A138T, T140I, C141R, C141G, C141A, C141Y, C141S, C141F, C141W, V143M, V143A, V143E, L145Q, W146C, W146L, L145R, V147G, P151T, P151A, P151S, P151H, P151R, P152S, P152R, P152L, T155P, T155A, V157F, R158H, R158L, A159V, A159P, A159S, A159D, A161T, A161D, Y163N, Y163H, Y163D, Y163S, Y163C, K164E, K164M, K164N, K164P, H168Y, H168P, H168R, H168L, H168Q, M169I, M169T, M169V, E171K, E171Q, E171G, E171A, E171V, E171D, V172D, V173M, V173L, V173G, R174W, R175G, R175C, R175H, C176R, C176G, C176Y, C176F, C176S, P177R, P177R, P177L, H178D, H178P, H178Q, H179Y, H179R, H179Q, R181C, R181Y, D186G, G187S, P190L, P190T, H193N, H193P, H193L, H193R, L194F, L194R, I195F, I195N, I195T, R196P, V197L, G199V, Y205N, Y205C, Y205H, D208V, R213Q, R213P, R213L, R213Q, H214D, H214R, S215G, S215I, S215R, V216M, V217G, Y220N, Y220H, Y220S, Y220C, E224D, I232F, I232N, I232T, I232S, Y234N, Y234H, Y234S, Y234C, Y236N, Y236H, Y236C, M237V, M237K, M237I, C238R, C238G, C238Y, C238W, N239T, N239S, S241Y, S241C, S241F, C242G, C242Y, C242S, C242F, G244S, G244C, G244D, G245S, G245R, G245C, | NM_001126112 |

| | | |
|---|---|---|
| | G245D, G245A, G245V, G245S, M246V, M246K, M246R, M246I, N247I, R248W, R248G, R248Q, R249G, R249W, R249T, R249M, P250L, I251N, L252P, I254S, I255F, I255N, I255S, L257Q, L257P, E258K, E258Q, D259Y, S261T, G262D, G262V, L265P, G266R, G266E, G266V, R267W, R267Q, R267P, E271K, V272M, V272L, R273S, R273G, R273C, R273H, R273P, R273L, V274F, V274D, V274A, V274G, V274L, C275Y, C275S, C275F, A276P, C277F, C277Y, P278T, P278A, P278S, P278H, P278R, P278L, G279E, R280G, R280K, R280T, R280I, R280S, D281N, D281H, D281Y, D281G, D281E, R282G, R282W, R282Q, R282P, E285K, E285V, E286G, E286V, E286K, K320N, L330R, G334V, R337C, R337L, A347T, L348F, T377P | |
| *U2AF1* | D14G, S34F, S34Y, R35L, R156H, R156Q, Q157R, Q157P | NM_006758 |
| *U2AF2* | R18W, Q143L, M144I, L187V, Q190L | NM_007279 |
| *WT1* | Frameshift/nonsense/splice-site | NM_024426 |
| *ZRSR2* | Frameshift/nonsense, R126P, E133G, C181F, H191Y, I202N, F239V, F239Y, N261Y, C280R, C302R, C326R, H330R, N382K | NM_005089 |

**Supplementary Table 3. CHIP variants ascertained through genotyping in the UK Biobank.**

| Variant | Gene | Chr | Location | Reference Allele | Effect Allele | Consequence |
|---|---|---|---|---|---|---|
| rs373145711 | *ASXL1* | 20 | 31021211 | C | T | Arg404Ter |
| rs373873045 | *DNMT3A* | 2 | 25459834 | C | A | Glu817Ter |
| rs765045799 | *DNMT3A* | 2 | 25462086 | T | C | Splice Acceptor |
| rs1190050788 | *DNMT3A* | 2 | 25466765 | A | C | Splice Donor |
| rs568207978 | *DNMT3A* | 2 | 25467083 | G | A | Arg598Ter |
| rs369109129 | *DNMT3A* | 2 | 25469922 | G | A | Gln374Ter |
| rs776841024 | *DNMT3A* | 2 | 25470556 | C | T | Trp306Ter |
| rs767439400 | *DNMT3A* | 2 | 25471030 | GGGCT | G | SerPro243Fs |
| rs77375493 | *JAK2* | 9 | 5073770 | G | T | Val617Phe |
| rs370735654 | *TET2* | 4 | 106196213 | C | T | Arg1516Ter |
| rs757144251 | *TET2* | 4 | 106157271 | TA | T | Lys725Fs |

Chr, chromosome; Fs, frameshift

**Supplementary Table 4. Association of CHIP with known risk factors across cohorts.**

| Risk Factor | FHS, n=4,240 | ARIC, n=7,414 | UK Biobank WES, n=201,409 | UK Biobank Array, n=239,316 |
|---|---|---|---|---|
| Age, year | OR 1.04 CI 1.03, 1.05, p<0.001 | OR 1.08 CI 1.06, 1.10, p<0.001 | OR 1.05 CI 1.04, 1.05, p<0.001 | OR 1.04 CI 1.02, 1.06, p<0.001 |
| Sex, female versus male | OR 1.24 CI 0.98, 1.56, p=0.07 | OR 1.03 CI 0.85, 1.25, p=0.77 | OR 1.06 CI 1.00, 1.11, p=0.04 | OR 1.37 CI 1.02, 1.74, p=0.01 |
| Current Smoking | OR 1.31 CI 0.98, 1.78, p=0.07 | OR 1.25 CI 1.00, 1.55, p=0.05 | OR 1.34 CI 1.21, 1.48, p<0.001 | OR 1.13 CI 0.74, 1.74, p=0.57 |
| Body mass index, kg/m$^2$ | OR 0.96 CI 0.94, 0.99, p=0.002 | OR 0.99 CI 0.97, 1.01, p=0.47 | OR 1.00 CI 1.00, 1.01 p=0.27 | OR 0.99 CI 0.97, 1.02 p=0.55 |
| Type 2 Diabetes mellitus | OR 0.96 CI 0.55, 1.69, p=0.90 | OR 0.96 CI 0.68, 1.35, p=0.81 | OR 0.92 CI 0.83, 1.03, p=0.14 | OR 0.89 CI 0.51, 1.54, p=0.67 |

Estimates were derived using multivariable logistic regression. ARIC, Atherosclerosis Risk in Communities study; CHIP; clonal hematopoiesis of indeterminate potential; CI, 95% confidence interval; FHS, Framingham Heart Study; OR, odds ratio; UKBB, UK Biobank; WES, whole exome sequencing.

**Supplementary Table 5. Mendelian randomization analyses of CHIP associations with chronic liver disease.**

| Mendelian randomization analysis | P-value threshold to select variants | Number of Variants | OR (95% CI) | p-value |
|---|---|---|---|---|
| Observational | NA | NA | 2.59 (1.65, 4.05) | $3x10^{-5}$ |
| MR-RAPS, primary analysis | p<0.0001 | 184 | 2.37 (1.57, 3.6) | $4x10^{-5}$ |
| MR-RAPS | p<0.001 | 824 | 2.05 (1.44, 2.91) | $6x10^{-5}$ |
| MR-RAPS | p<0.00005 | 90 | 2.34 (0.97, 5.67) | 0.06 |
| IVW | p<0.0001 | 184 | 2.44 (1.29, 4.61) | 0.007 |
| MR-Egger | p<0.0001 | 184 | 20.4 (4.04, 103) | 0.0004 |
| MR-PRESSO | p<0.0001 | 184 | 2.45 (1.3, 4.59) | 0.006 |
| Multivariate IVW | p<0.0001 | 184 | 2.56 (1.41, 4.63) | 0.002 |

Number of variants refers to the number of SNPs used in the Mendelian randomization score. CHIP; clonal hematopoiesis of indeterminate potential; CI, confidence interval; MR-RAPS, Mendelian randomization with adjusted profile score; IVW, inverse variance weighted regression; MR-PRESSO, Mendelian randomization Pleiotropy RESidual Sum and Outlier; OR, odds ratio.

**Supplementary Table 6. Genetic variants used for Mendelian randomization analyses.**

https://docs.google.com/spreadsheets/d/1NHGfy2fD7rDUEog2G9Y6HuX_gmVJ5YU9cE0Nki5lhI8/edit?usp=sharing

**Supplementary Table 7. Histologic grading criteria for steatohepatitis in mice.**

| | |
|---|---|
| ***Steatosis*** | |
| Grade 0 | Absent |
| Grade 1 | < 33% of hepatocytes |
| Grade 2 | 33-66% of hepatocytes |
| Grade 3 | > 66% of hepatocytes |
| ***Inflammation*** | |
| Grade 0 | Absent |
| Grade 1 | < 100 inflammatory cells per focus or < 3 inflammatory foci per 20x field |
| Grade 2 | 100-500 inflammatory cells per focus or 3 - 4 inflammatory foci per 20x field |
| Grade 3 | > 500 inflammatory cells per focus or > 5 inflammatory foci per 20x field |
| ***Hepatocyte Injury/Ballooning*** | |
| Grade 0 | Absent |
| Grade 1 | Rare balloon cells |
| Grade 2 | Widespread hepatocyte ballooning and apoptosis |
| ***Modified NAFLD activity score (NAS)*** | |
| Grade 1 | 0-2 points |
| Grade 2 | 3-4 points |
| Grade 3 | > 5 points |

Modified from NASH Clinical Research Network scoring system for NAFLD (Kleiner et al., 2005). NAFLD, nonalcoholic fatty liver disease; NASH, nonalcoholic steatohepatitis.

**Supplementary Table 8. Differentially regulated genes in sorted liver macrophages from *Tet2*$^{-/-}$ and control bone marrow transplanted mice**.

https://docs.google.com/spreadsheets/d/1Q3pFPsMj1IdCYoXc578pMAQiWfy-C-uc5ZeZRwsnn_E/edit?usp=sharing

**Supplementary Table 9. Gene set enrichment analysis of *Tet2*$^{-/-}$ versus control sorted liver macrophages.**

https://docs.google.com/spreadsheets/d/1TUambdLrHXdhv0Ios8jo6ZWN6XunVuci1UmT32vcKoQ/edit?usp=sharing

**Supplementary Table 10. Differentially regulated genes in bulk liver transcripts from *Tet2*$^{-/-}$ and control bone marrow transplanted mice.**

https://docs.google.com/spreadsheets/d/1I_dT0dFkaokqKcwASyefYz2sBk7V3_ZK1dUSabwAzmM/edit?usp=sharing

**Supplementary Table 11. Gene set enrichment analysis of *Tet2*$^{-/-}$ versus control bulk liver RNA.**

https://docs.google.com/spreadsheets/d/1YG66y5MZeemzcsKdKAszCyWUP3ngPOxVoiQe4XX03Cs/edit?usp=sharing

**Supplementary Figure 1. Study design of chronic liver disease associations with CHIP.**

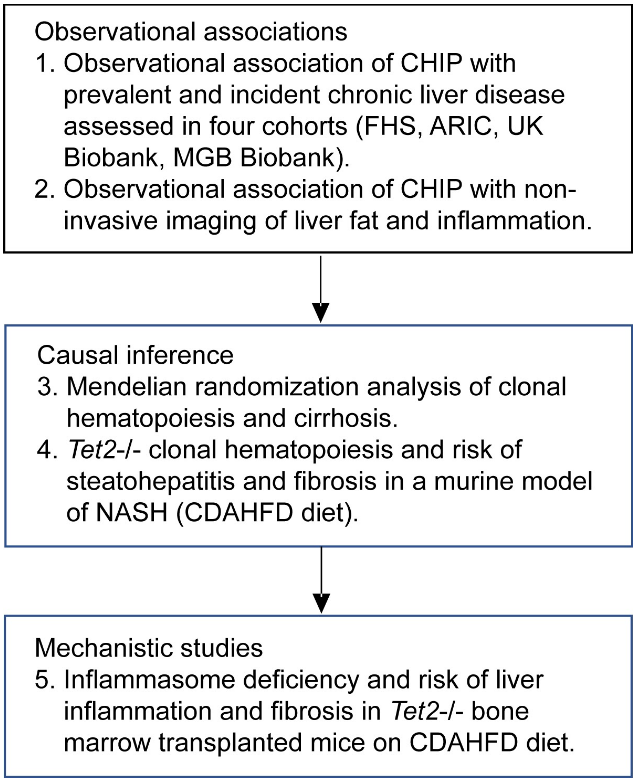


ARIC, Atherosclerosis Risk in Communities study; CDAHFD, choline deficient amino acid defined high fat diet; CHIP, clonal hematopoiesis of indeterminate potential; FHS, Framingham Heart Study; MGB, Mass General Brigham; NASH, nonalcoholic steatohepatitis.

**Supplementary Figure 2. Proportion of CHIP by mutated gene among individuals with CHIP.**

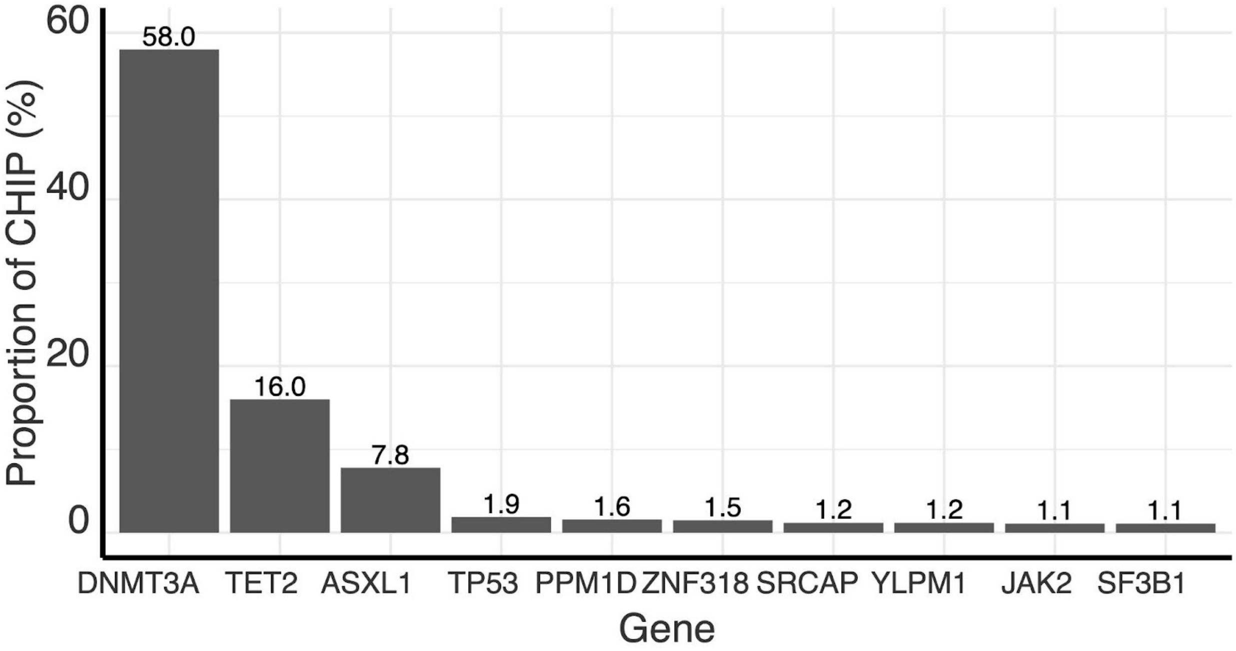


CHIP, clonal hematopoiesis of indeterminate potential.

**Supplementary Figure 3. Prevalence of CHIP by age**.

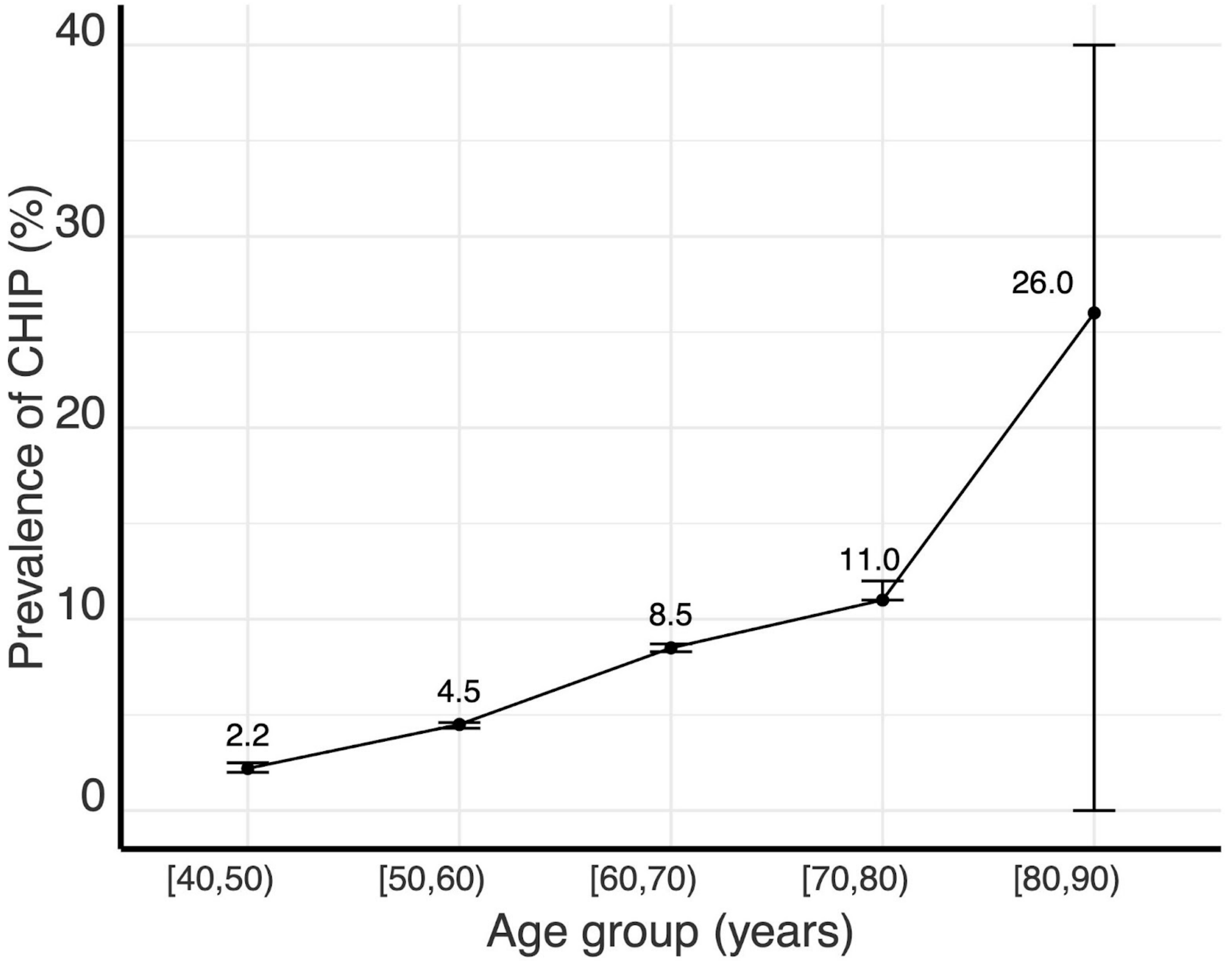


CHIP, clonal hematopoiesis of indeterminate potential.

**Supplementary Figure 4. Association of CHIP with prevalent chronic liver disease by variant allele fraction**.

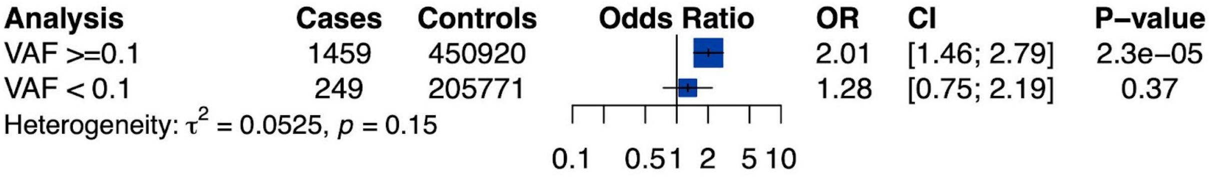


Estimates derived using logistic regression with adjustment for age and sex in the UK Biobank, Framingham Heart Study and Atherosclerosis Risk in Communities study and pooled using inverse variance weighted fixed effects meta-analysis. CHIP, clonal hematopoiesis of indeterminate potential; CI, 95% confidence interval; OR, odds ratio; VAF, variant allele fraction

**Supplementary Figure 5. Association of CHIP with chronic liver disease by mutated gene**.

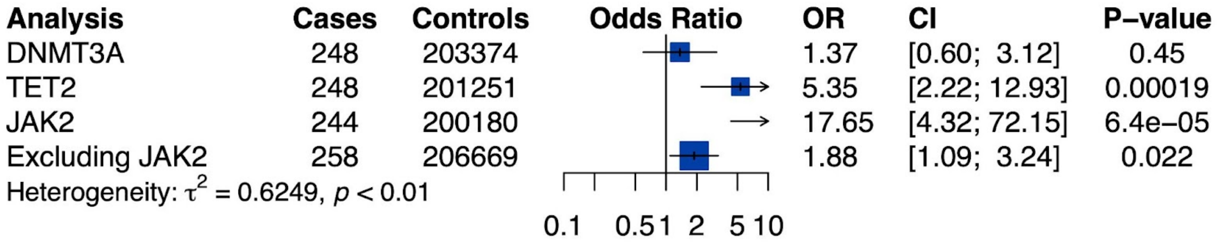


Estimates derived in Framingham, ARIC and the UK BIobank using logistic regression adjusted for age and sex. Estimates pooled across cohorts using inverse variance weighted fixed effects meta-analysis. CHIP, clonal hematopoiesis of indeterminate potential; CI, 95% confidence interval; OR, odds ratio.

**Supplementary Figure 6. Cumulative risk of chronic liver disease by clonal hematopoiesis status in the UK Biobank**.

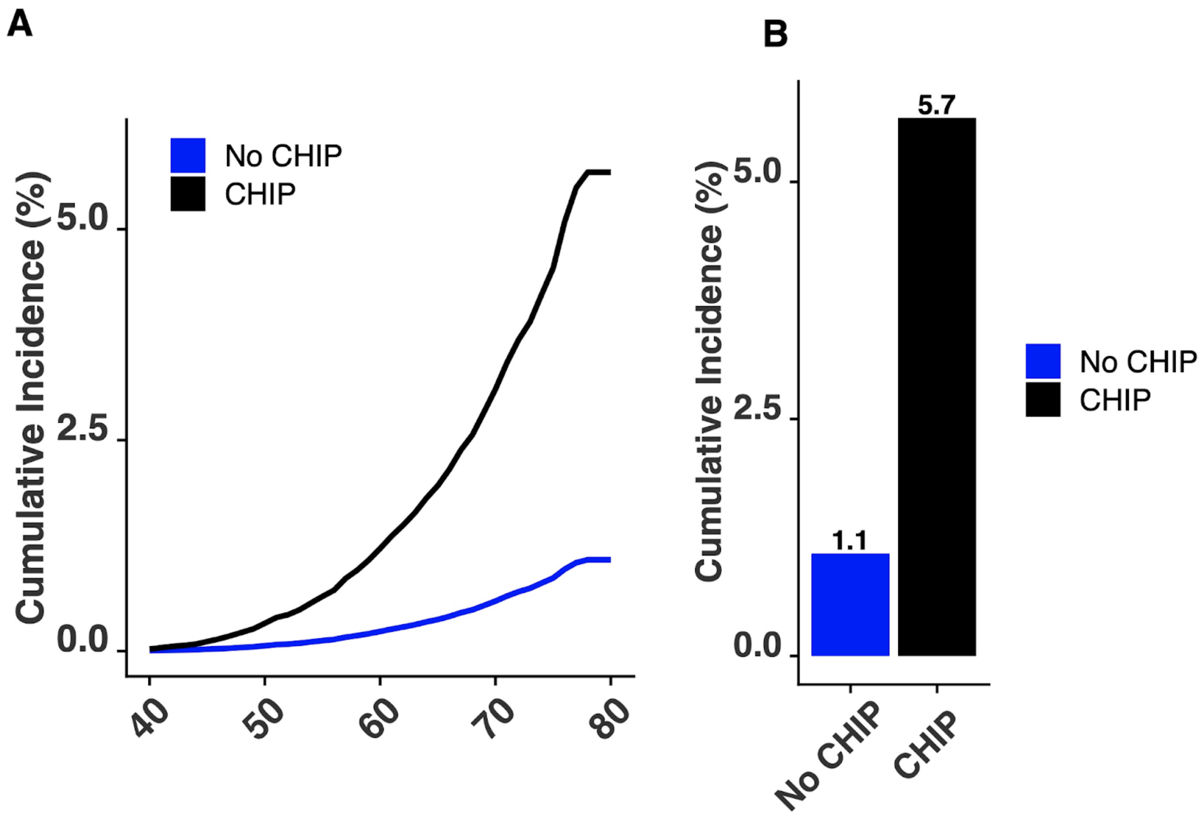


Cumulative risk of chronic liver disease by age was modeled using Cox proportional hazards model with age as the underlying time variable and adjustment for sex. CHIP, clonal hematopoiesis of indeterminate potential.

**Supplementary Figure 7. Association of CHIP with serum biomarkers in the UK Biobank**.

| Biomarker | N | Beta (SD) | 95% CI | Clinical Units | P−value |
|---|---|---|---|---|---|
| **LFTs** | | | | | |
| ALT | 388905 | −0.01 | [−0.05; 0.03] | −0.0045 (−0.021, 0.012) U/L | 0.6 |
| AST | 387607 | −0.00 | [−0.04; 0.04] | −0.00026 (−0.01, 0.0099) U/L | 0.96 |
| ALP | 388791 | 0.05 | [ 0.02; 0.09] | 0.015 (0.005, 0.026) U/L | 0.0036 |
| GGT | 386238 | 0.03 | [−0.01; 0.07] | 1.2 (−0.43, 2.8) U/L | 0.15 |
| TBili | 384783 | 0.07 | [ 0.03; 0.11] | 0.31 (0.15, 0.48) mg/dl | 0.00016 |
| **Inflammation** | | | | | |
| CRP | 386859 | 0.04 | [ 0.00; 0.08] | 0.19 (0.018, 0.35) mg/dl | 0.03 |
| **CBC** | | | | | |
| WBC | 393124 | 0.08 | [ 0.04; 0.12] | 0.16 (0.083, 0.24) /uL | 4.6e−05 |
| Hemoglobin | 393128 | 0.02 | [−0.01; 0.05] | 0.022 (−0.016, 0.06) g/dl | 0.26 |
| Platelet | 393127 | 0.14 | [ 0.10; 0.18] | 8.5 (6.3, 11) 10^9/L | <0.001 |

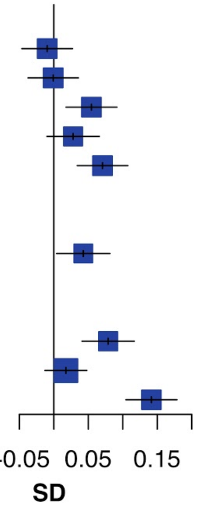


ALP, alkaline phosphatase; ALT, alanine transaminase; AST, aspartate transaminase; CBC, complete blood count; CHIP, clonal hematopoiesis of indeterminate potential; CI, confidence interval; CRP, C-reactive protein; GGT, gamma-glutamyl transferase; LFT, liver function test; TBili, total bilirubin; WBC, white blood cell count. A p-value of 0.006 after Bonferroni adjustment (0.05/9 = 0.006) was considered significant.

**Supplementary Figure 8. Association of CHIP with serum biomarkers in the UK Biobank with carriers of *JAK2*-mutant CHIP excluded.**

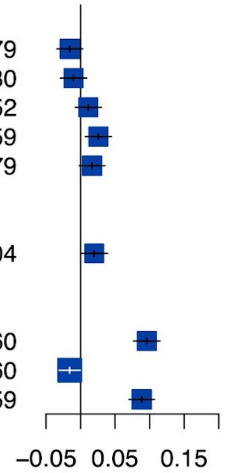


| Biomarker | N | Beta (SD) | 95% CI | Clinical Units | P−value |
|---|---|---|---|---|---|
| **LFTs** | | | | | |
| ALT | 190279 | −0.02 | [−0.03; 0.00] | −0.0071 (−0.016, 0.0014) U/L | 0.1 |
| AST | 189680 | −0.01 | [−0.03; 0.01] | −0.0028 (−0.008, 0.0025) U/L | 0.3 |
| ALP | 190352 | 0.01 | [−0.01; 0.03] | 0.0031 (−0.0022, 0.0084) U/L | 0.25 |
| GGT | 190259 | 0.03 | [ 0.01; 0.04] | 1.1 (0.28, 1.8) U/L | 0.008 |
| TBili | 189579 | 0.02 | [ 0.00; 0.04] | 0.074 (−0.0093, 0.16) mg/dl | 0.082 |
| **Inflammation** | | | | | |
| CRP | 189904 | 0.02 | [ 0.00; 0.04] | 0.085 (0.0023, 0.17) mg/dl | 0.044 |
| **CBC** | | | | | |
| WBC | 194160 | 0.10 | [ 0.08; 0.12] | 0.19 (0.16, 0.23) /uL | 7.9e−23 |
| Hemoglobin | 194160 | −0.02 | [−0.03; 0.00] | −0.02 (−0.038, −0.00064) g/dl | 0.043 |
| Platelet | 194159 | 0.09 | [ 0.07; 0.11] | 5.3 (4.2, 6.4) 10^9/L | 8e−21 |

ALP, alkaline phosphatase; ALT, alanine transaminase; AST, aspartate transaminase; CBC, complete blood count; CHIP, clonal hematopoiesis of indeterminate potential; CI, confidence interval; CRP, C-reactive protein; GGT, gamma-glutamyl transferase; LFT, liver function test; TBili, total bilirubin; WBC, white blood cell count. A p-value of 0.006 after Bonferroni adjustment (0.05/9 = 0.006) was considered significant.

**Supplementary Figure 9. Mendelian randomization analysis of CHIP association with chronic liver disease.** (**A)** Effect of genetic variants against exposure (CHIP) and outcome (cirrhosis). **(B)** Mendelian randomization estimates of the association of CHIP with chronic liver disease.

## A

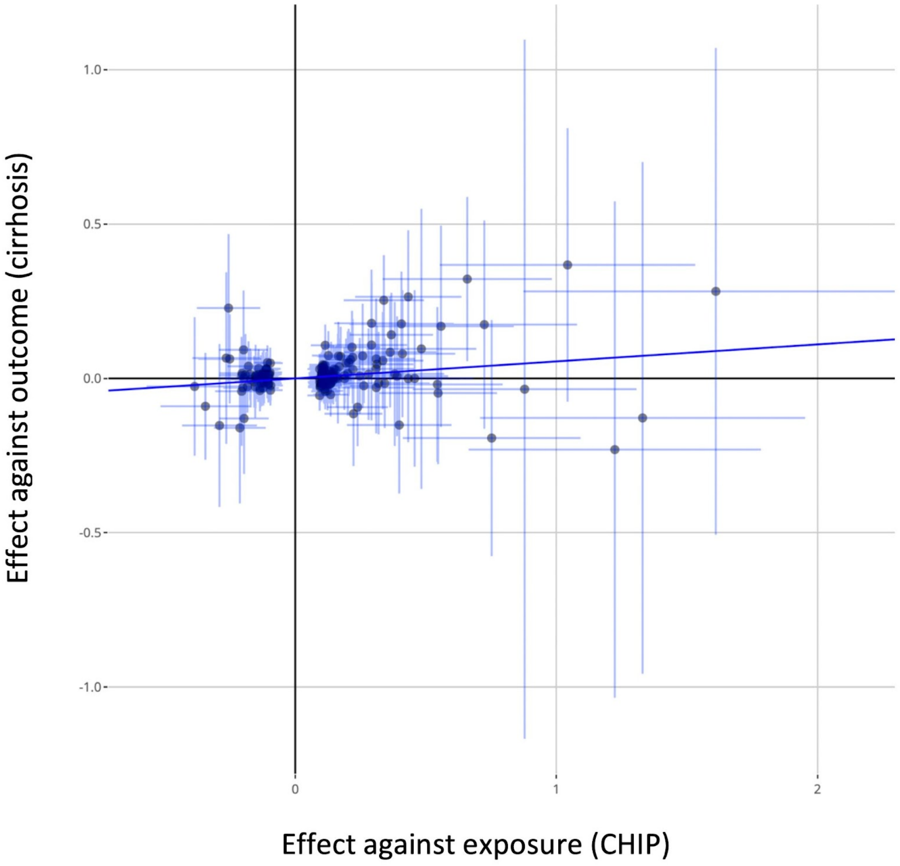


## B

| Analysis | Controls | Cases | Odds Ratio | OR | CI | P–value |
|---|---|---|---|---|---|---|
| Observational | 421238 | 1310 | | 2.59 | [1.65; 4.05] | $3.2x10^{-5}$ |
| Mendelian Randomization | 487780 | 5777 | | 2.37 | [1.57; 3.60] | $4.5x10^{-5}$ |

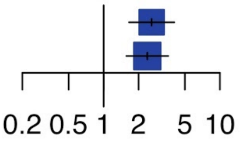


Estimates were derived using MR-RAPS (Mendelian Randomization using Robust Adjusted Profile Score) with 184 independent genetic variants with significance of p<0.0001. CHIP, clonal hematopoiesis of indeterminate potential; CI, 95% confidence interval; OR, odds ratio.

**Supplementary Figure 10. Liver and hematologic parameters of *Ldlr-/-* (A-F) and B6.SJL (G-L) mice transplanted with *Tet2-/-* or control bone marrow cells and fed Western diet for 10 weeks or CDAHFD for 11 weeks, respectively.**

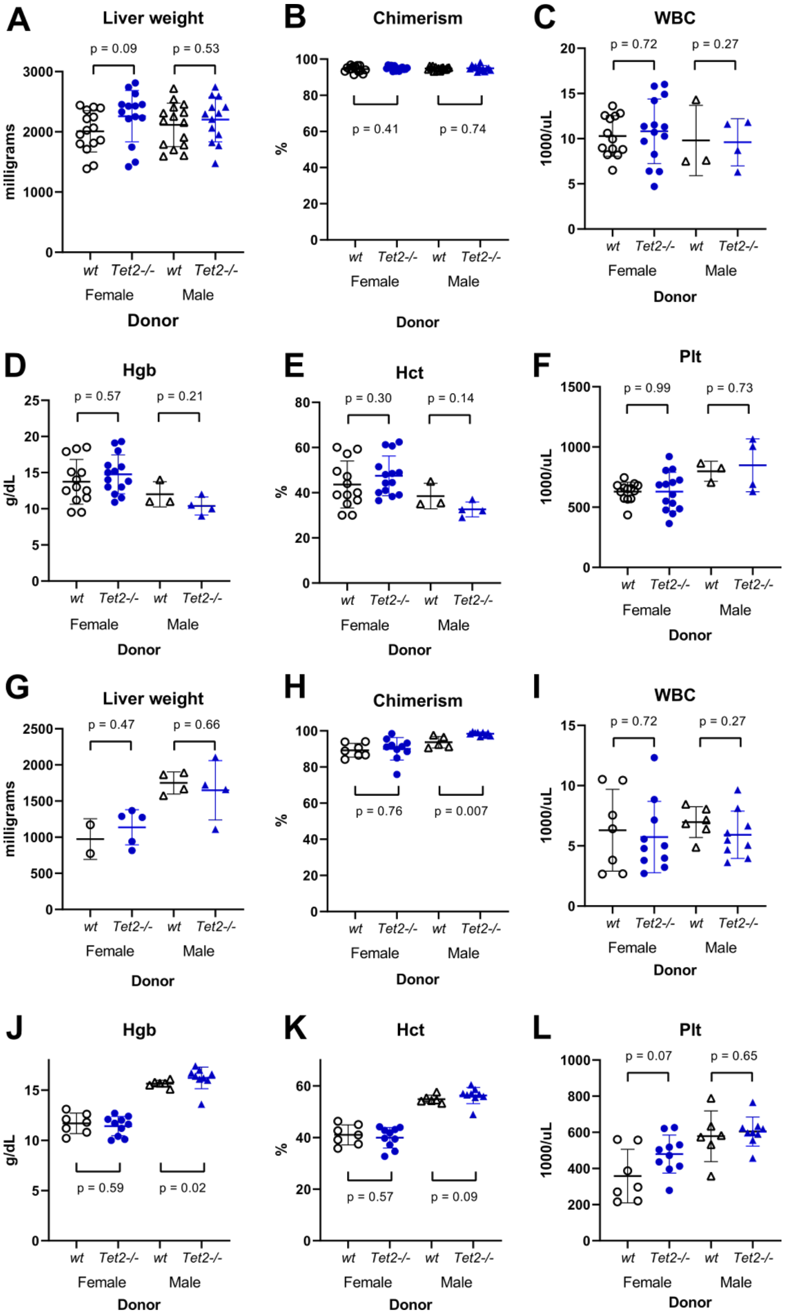


CDAHFD, choline deficient amino acid defined high fat diet; Hct, hematocrit; Hgb, hemoglobin concentration; Plt, platelet count; WBC, white blood cell count.

**Supplementary Figure 11. Steatohepatitis and liver fibrosis in *Tet2*$^{-/-}$ and *Dnmt3a*$^{-/-}$ transplanted mice. (A)** Histologic features of steatohepatitis in *Ldlr*$^{-/-}$ mice transplanted with *Tet2*$^{-/-}$ (n=25) and control (n=20) bone marrow and fed Western diet for 10 weeks were graded on a semiquantitative scale and aggregated into a NAFLD activity score (NAS) using CRN histologic scoring criteria. **(B-E)** Graded histologic features included steatosis (B), inflammatory foci (C), hepatocyte ballooning injury (D, arrowhead) and apoptosis (E, arrowhead). **(F)** Masson's trichrome staining demonstrates absence of perivenular fibrosis in control and *Tet2*$^{-/-}$ transplanted mice. **(G)** B6.SJL mice were transplanted with *Dnmt3a*$^{-/-}$ (n=24) or control (n=19) bone marrow cells and fed CDAHFD for 11 weeks. Steatohepatitis was assessed histologically for steatosis, inflammation, and hepatocyte ballooning injury. Collagen fibrosis was measured by Masson's trichrome staining. **(H)** *B6.SJL* mice were transplanted with *Tet2*$^{-/-}$ (n=21) or control (n=15) bone marrow cells and fed CDAHFD for 11 weeks, then switched to regular diet for 10 days. Compared to control animals, *Tet2*$^{-/-}$ transplanted mice show similar resolution of liver fat but show persistently greater inflammation and more hepatocyte ballooning injury. Collagen fibrosis, as measured by Masson's trichrome staining, was not significantly different.

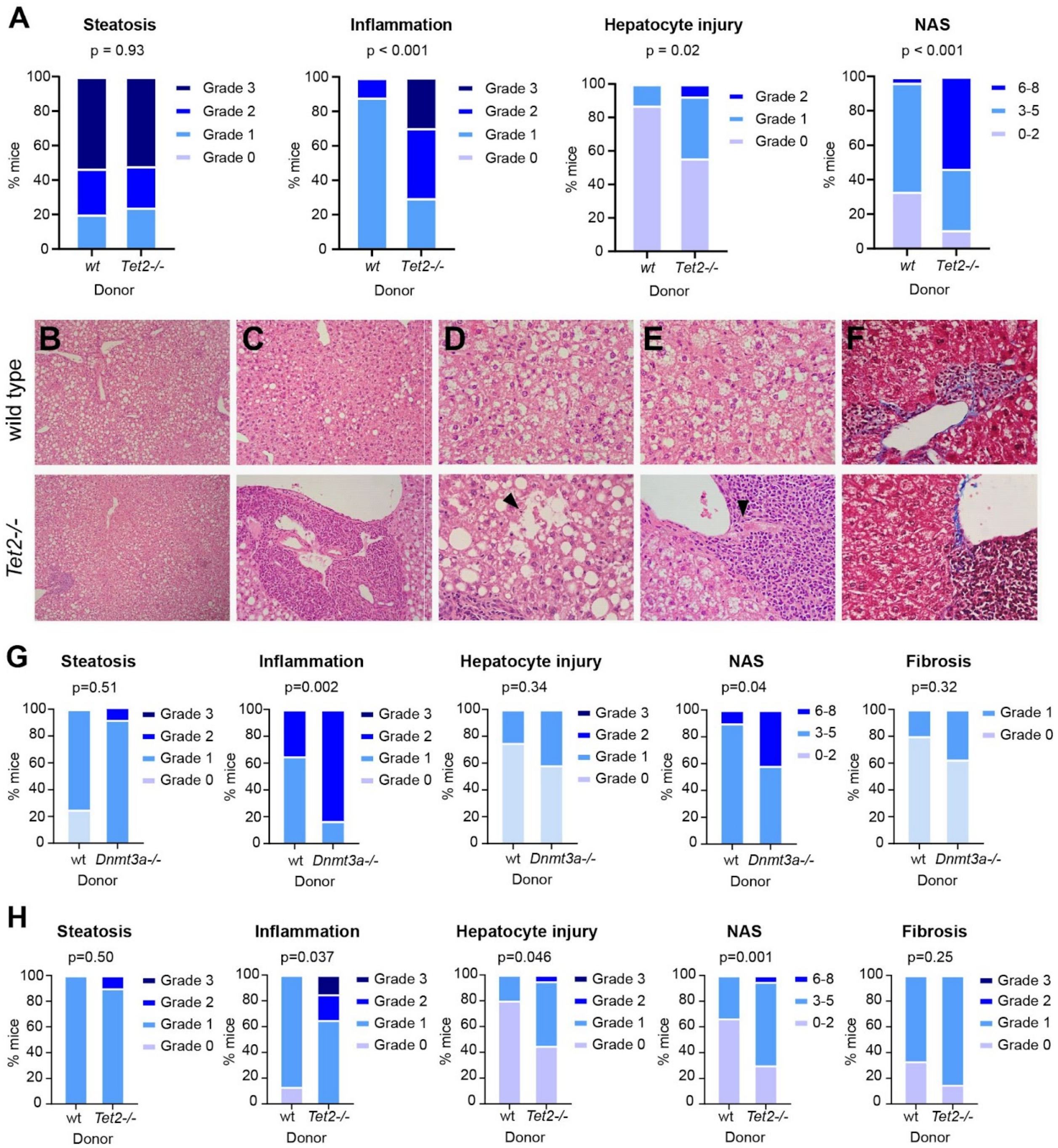
A
Steatosis
p = 0.93
Inflammation
p < 0.001
Hepatocyte injury
p = 0.02
NAS
p < 0.001
% mice
Grade 3
Grade 2
Grade 1
Grade 0
6-8
3-5
0-2
wt
Tet2-/-
Donor
B
C
D
E
F
wild type
Tet2-/-
G
Steatosis
p=0.51
Inflammation
p=0.002
Hepatocyte injury
p=0.34
NAS
p=0.04
Fibrosis
p=0.32
wt
Dnmt3a-/-
Donor
H
Steatosis
p=0.50
Inflammation
p=0.037
Hepatocyte injury
p=0.046
NAS
p=0.001
Fibrosis
p=0.25
wt
Tet2-/-
Donor

**Supplementary Figure 12. Gene expression analysis of sorted liver macrophages and bulk liver RNA in *Tet2*$^{-/-}$ transplanted mice fed CDAHFD. (A)** Immunohistochemical stains demonstrate the presence of CD45.2$^{+}$CD45.1$^{-}$F4/80$^{+}$ macrophages in livers of bone marrow transplanted mice fed CDAHFD. **(B)** Selected enriched gene sets in sorted *Tet2*$^{-/-}$ liver macrophages from transplanted mice fed CDAHFD. The most highly enriched genes in each signature are shown. **(C)** Selected enriched gene sets in bulk livers of *Tet2*$^{-/-}$ transplanted mice fed CDAHFD. The most highly enriched genes in each signature are shown. NES, normalized enrichment score.

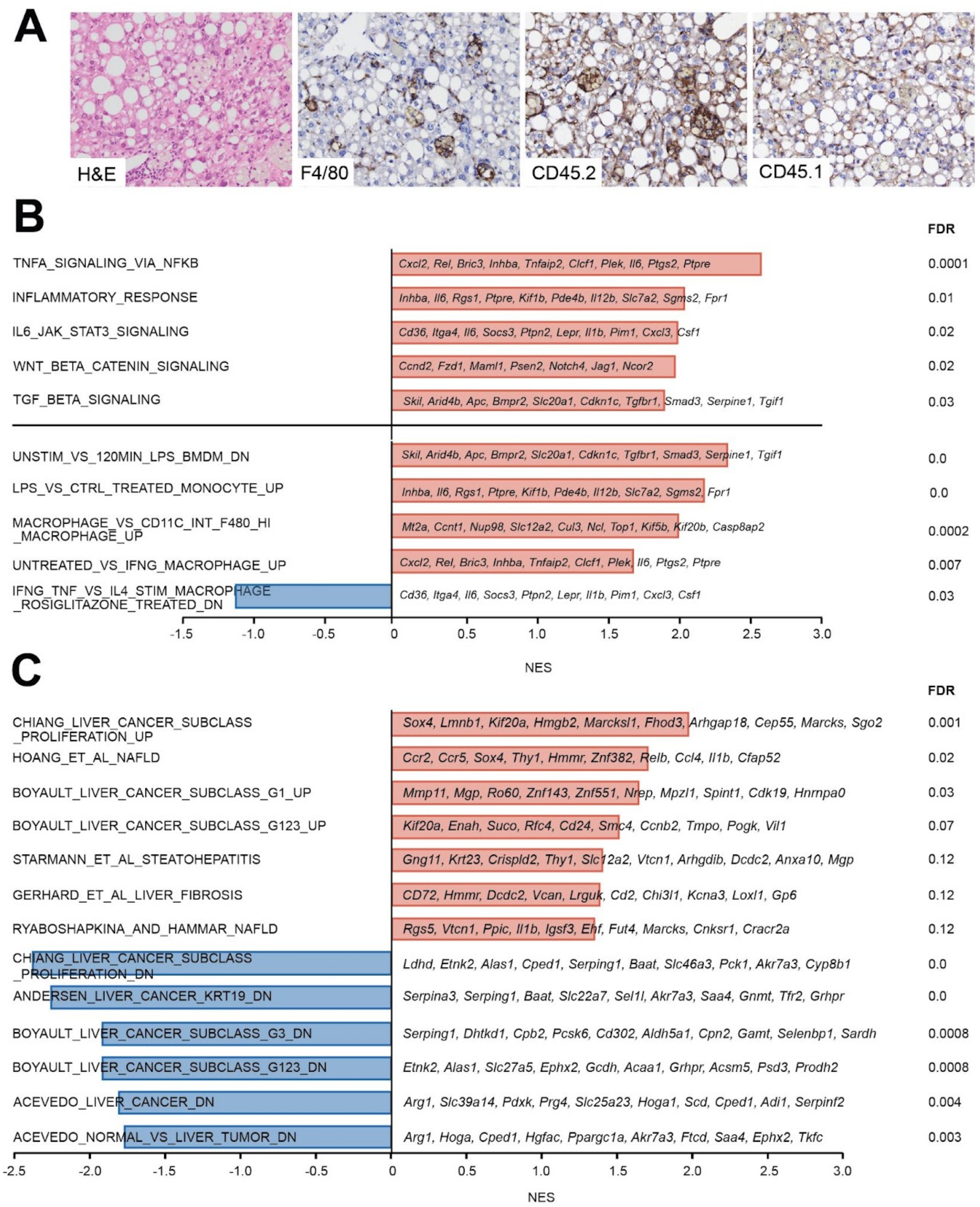
A
H&E
F4/80
CD45.2
CD45.1
B
FDR
TNFA_SIGNALING_VIA_NFKB
Cxcl2, Rel, Bric3, Inhba, Tnfaip2, Clcf1, Plek, Il6, Ptgs2, Ptpre
0.0001
INFLAMMATORY_RESPONSE
Inhba, Il6, Rgs1, Ptpre, Kif1b, Pde4b, Il12b, Slc7a2, Sgms2, Fpr1
0.01
IL6_JAK_STAT3_SIGNALING
Cd36, Itga4, Il6, Socs3, Ptpn2, Lepr, Il1b, Pim1, Cxcl3, Csf1
0.02
WNT_BETA_CATENIN_SIGNALING
Ccnd2, Fzd1, Maml1, Psen2, Notch4, Jag1, Ncor2
0.02
TGF_BETA_SIGNALING
Skil, Arid4b, Apc, Bmpr2, Slc20a1, Cdkn1c, Tgfbr1, Smad3, Serpine1, Tgif1
0.03
UNSTIM_VS_120MIN_LPS_BMDM_DN
Skil, Arid4b, Apc, Bmpr2, Slc20a1, Cdkn1c, Tgfbr1, Smad3, Serpine1, Tgif1
0.0
LPS_VS_CTRL_TREATED_MONOCYTE_UP
Inhba, Il6, Rgs1, Ptpre, Kif1b, Pde4b, Il12b, Slc7a2, Sgms2, Fpr1
0.0
MACROPHAGE_VS_CD11C_INT_F480_HI_MACROPHAGE_UP
Mt2a, Ccnt1, Nup98, Slc12a2, Cul3, Ncl, Top1, Kif5b, Kif20b, Casp8ap2
0.0002
UNTREATED_VS_IFNG_MACROPHAGE_UP
Cxcl2, Rel, Bric3, Inhba, Tnfaip2, Clcf1, Plek, Il6, Ptgs2, Ptpre
0.007
IFNG_TNF_VS_IL4_STIM_MACROPHAGE_ROSIGLITAZONE_TREATED_DN
Cd36, Itga4, Il6, Socs3, Ptpn2, Lepr, Il1b, Pim1, Cxcl3, Csf1
0.03
-1.5
-1.0
-0.5
0
0.5
1.0
1.5
2.0
2.5
3.0
NES
C
FDR
CHIANG_LIVER_CANCER_SUBCLASS_PROLIFERATION_UP
Sox4, Lmnb1, Kif20a, Hmgb2, Marcksl1, Fhod3, Arhgap18, Cep55, Marcks, Sgo2
0.001
HOANG_ET_AL_NAFLD
Ccr2, Ccr5, Sox4, Thy1, Hmmr, Znf382, Relb, Ccl4, Il1b, Cfap52
0.02
BOYAULT_LIVER_CANCER_SUBCLASS_G1_UP
Mmp11, Mgp, Ro60, Znf143, Znf551, Nrep, Mpzl1, Spint1, Cdk19, Hnrnpa0
0.03
BOYAULT_LIVER_CANCER_SUBCLASS_G123_UP
Kif20a, Enah, Suco, Rfc4, Cd24, Smc4, Ccnb2, Tmpo, Pogk, Vil1
0.07
STARMANN_ET_AL_STEATOHEPATITIS
Gng11, Krt23, Crispld2, Thy1, Slc12a2, Vtcn1, Arhgdib, Dcdc2, Anxa10, Mgp
0.12
GERHARD_ET_AL_LIVER_FIBROSIS
CD72, Hmmr, Dcdc2, Vcan, Lrguk, Cd2, Chi3l1, Kcna3, Loxl1, Gp6
0.12
RYABOSHAPKINA_AND_HAMMAR_NAFLD
Rgs5, Vtcn1, Ppic, Il1b, Igsf3, Ehf, Fut4, Marcks, Cnksr1, Cracr2a
0.12
CHIANG_LIVER_CANCER_SUBCLASS_PROLIFERATION_DN
Ldhd, Etnk2, Alas1, Cped1, Serping1, Baat, Slc46a3, Pck1, Akr7a3, Cyp8b1
0.0
ANDERSEN_LIVER_CANCER_KRT19_DN
Serpina3, Serping1, Baat, Slc22a7, Sel1l, Akr7a3, Saa4, Gnmt, Tfr2, Grhpr
0.0
BOYAULT_LIVER_CANCER_SUBCLASS_G3_DN
Serping1, Dhtkd1, Cpb2, Pcsk6, Cd302, Aldh5a1, Cpn2, Gamt, Selenbp1, Sardh
0.0008
BOYAULT_LIVER_CANCER_SUBCLASS_G123_DN
Etnk2, Alas1, Slc27a5, Ephx2, Gcdh, Acaa1, Grhpr, Acsm5, Psd3, Prodh2
0.0008
ACEVEDO_LIVER_CANCER_DN
Arg1, Slc39a14, Pdxk, Prg4, Slc25a23, Hoga1, Scd, Cped1, Adi1, Serpinf2
0.004
ACEVEDO_NORMAL_VS_LIVER_TUMOR_DN
Arg1, Hoga, Cped1, Hgfac, Ppargc1a, Akr7a3, Ftcd, Saa4, Ephx2, Tkfc
0.003
-2.5
-2.0
-1.5
-1.0
-0.5
0
0.5
1.0
1.5
2.0
2.5
3.0
NES

### Extended Materials and Methods

*Study samples*

We examined whether CHIP is associated with elevated risk of chronic liver disease using five datasets. For prevalent chronic liver disease, we tested the association of CHIP with chronic liver disease using data from the Framingham Heart Study (FHS, n=4230) and Atherosclerosis Risk in Communities study (ARIC, n=7414). For incident chronic liver disease, we tested the association of CHIP ascertained through whole exome sequencing using a subsample of the UK Biobank who underwent whole exome sequencing (n=201,409) and a separate subsample of UK Biobank who underwent array based genotyping using genotyped CHIP variants (n=239,316). Only a subset of CHIP variants were available on the genotyped UK Biobank array (**Supp. Table 3**). Individuals with prevalent leukemia or other hematologic malignancy were excluded from analysis. All patients with whole exome sequencing data from FHS, ARIC and UKBB were included in the analysis. Median duration of follow up for incident disease analysis was 8.1 years.

We defined chronic liver disease as the development of liver fibrosis or cirrhosis, combining the following ICD10 diagnostic codes: K70.2 (alcoholic fibrosis and sclerosis of the liver), K70.3 (alcoholic cirrhosis of the liver), K70.4 (alcoholic hepatic failure), K74.0 (hepatic fibrosis), K74.1 (hepatic sclerosis), K74.2 (hepatic fibrosis with hepatic sclerosis), K74.6 (other and unspecified cirrhosis of liver), K76.6 (portal hypertension), or I85 (esophageal varices). These ICD10 codes have previously been demonstrated to have high specificity for identification of patients with cirrhosis compared to physician review and associate strongly with known cirrhosis loci.[1]

In addition to examining all-cause chronic liver disease, we also examined the association of CHIP with subtypes of chronic liver disease. Non-alcoholic fatty liver disease was defined as chronic liver disease among individuals consuming less than 21 drinks per week for men and less than 14 drinks per week for women and no history of hepatitis B or hepatitis C as per the American Association for the Study of Liver Diseases guidelines.[2] Alcohol-related liver disease was defined as chronic liver disease among individuals with excess alcohol intake (≥21 drinks per week for men or ≥14 drinks per week for women) and no history of hepatitis B or hepatitis C. Seven individuals with liver disease and a history of known hepatitis B or hepatitis C were excluded from the subtype of chronic liver disease analysis.

To examine whether CHIP associates with serum biomarkers or liver imaging, we used data from the UK Biobank. Blood samples were collected from all UK Biobank participants during their initial enrollment visit. These samples were used for both whole exome sequencing and for biomarker assays. In a cross-sectional analysis, we tested whether carrying CHIP associated with serum liver enzyme levels (alanine transaminase levels, aspartate transaminase levels, alkaline phosphatase levels, gamma glutamyl transferase levels) and serum inflammatory biomarkers (C-reactive protein, platelet count, hemoglobin and white blood cell count). Serum liver enzyme levels and C-reactive protein were measured by immunoassay using a Beckman Coulter AU5800. Blood cell counts were measured using a Beckman Coulter LH 750.

We examined whether CHIP status associated with liver fat and liver inflammation and fibrosis in 4434 individuals in the UK Biobank with whole exome sequence data who underwent Multiscan magnetic resonance imaging of their liver.[3] Liver fat was measured using the proton density fat fraction. Liver inflammation and fibrosis was measured using iron corrected T1 relaxation time (cT1).[3]

*Whole-exome sequencing and CHIP ascertainment*

We identified individuals with CHIP on the basis of a prespecified list of variants in 74 genes known to be recurrently mutated in myeloid cancers (Supplementary Table 1). For analyses of prevalent CHIP, CHIP status was ascertained using whole exome sequencing.[4,5] Whole exome sequences from 7414 individuals in the Atherosclerosis Risk in Communities study were obtained from dbGAP (Accession phs000280). DNA from ARIC was obtained from whole blood samples at the time of study enrollment. For Framingham Heart Study, exome sequencing was performed as part of the Trans-Omics for Precision Medicine program (TOPMED). In the UK Biobank, whole exome sequencing was performed centrally at the Regeneron Genetics Center. We analyzed 201,309 whole exomes from unrelated individuals in the UK Biobank. Individuals with prevalent leukemia or other hematologic malignancy were excluded from analysis.

CHIP was ascertained using whole exome sequencing as described in Bick A. et al., Nature 2020. Briefly, GATK Mutect2 somatic variant caller was applied to each sample from UK BIobank, ARIC and Framingham study to identify somatic variation. Mutect2 identifies sites where there is evidence of variation among reads. To exclude germline variants, we used an external reference comprised of gnomAD and a panel of normal derived from 100 randomly selected individuals under the age of 40 years. CHIP was then identified as somatic variation within a prespecified list of variants in 74 genes recurrently mutated in myeloid cancers (**Supp. Table 2**). CHIP was restricted to prespecified somatic variants in the 74 genes with variant allele fraction >2%.

In an analysis of incident chronic liver disease, we examined whether array-derived germline genotyped CHIP variants in *JAK2*, *ASXL1*, *DNMT3A* and *TET2* were associated with incident chronic liver disease in the UK Biobank. We examined genotyping fidelity of each variant using manual examination of imaging files with ScatterShot. Individuals who were included in the whole exome sequencing and those with prevalent liver disease were excluded from analysis.

*Mouse models*

*Ldlr*$^{-/-}$ and B6.SJL male and female mice at 8 weeks were exposed to 10 Gy total body irradiation and transplanted via retro-orbital injection with 1,000,000 to 2,000,000 bone marrow cells harvested from sex-matched *vavCre*$^{+}$*Tet2*$^{fl/-}$, *vavCre*$^{+}$*Tet2*$^{fl/-}$*Nlrp3*$^{-/-}$, and control *vavCre*$^{+}$

mice. After hematopoietic reconstitution was confirmed by peripheral blood analysis and flow cytometry at 4 weeks, mice were fed an atherogenic Western diet containing 0.2% cholesterol and 42 kcal % fat (TD.88137; Envigo) or a choline-deficient, L-amino acid defined, high-fat diet containing 60 kcal % fat and 0.1% methionine (CDAHFD, A06071302; Research Diets) for defined time periods. All animal experiments were approved by the Institutional Animal Care and Use Committee (IACUC) at Brigham and Women's Hospital and Dana Farber Cancer Institute.

*Mouse peripheral blood analysis*

Peripheral blood was collected from the retro-orbital sinus into EDTA tubes. Complete blood counts were obtained using a Hemavet 950 analyzer. After red cell lysis, cells were resuspended in PBS supplemented with 2% FBS for flow cytometric analysis using a BD FACScanto II analyzer. The following antibodies (all from Biolegend) were used for multicolor flow cytometry: mouse anti-mouse CD45.1 FITC (A20), mouse anti-mouse CD45.2 PE (104), rat anti-mouse CD3 PE-Cy7 (17A2), rat anti-human/mouse CD11b APC-Cy7 (M1/70), rat anti-mouse Gr-1 Pacific Blue (RB6-8C5), rat anti-human/mouse CD45R/B220 BV510 (RA3-6B2). Plasma was obtained by centrifugation at 1000 x g for 10 min at 4 °C and cytokine levels measured using Luminex-based mouse cytokine/chemokine magnetic bead panel (Millipore).

*Bone marrow derived macrophages*

Bone marrow cells from *vavCre*$^{+}$*Tet2*$^{fl/-}$, *vavCre*$^{+}$*Tet2*$^{fl/-}$*Nlrp3*$^{-/-}$, and control *vavCre*$^{+}$ mice were cultured with recombinant M-CSF (Preprotech) for 8 days and terminal differentiation confirmed using flow cytometry.  Macrophages were exposed to 10 ng/mL lipopolysaccharide (Sigma) for 2 hours and stimulated for 6 hours with palmitic acid (300 µM; Sigma) or cholesterol monohydrate crystals (200 µg/mL; Sigma) prepared as previously described[6]. Culture supernatant was centrifuged at 1000 x g for 10 min at 4 °C and cytokine levels measured using Luminex-based mouse cytokine/chemokine magnetic bead panel (Millipore).

*Liver histology*

Mouse livers were fixed in 10% formalin for 24 hours. Paraffin embedded tissue blocks were sectioned and stained using hematoxylin & eosin for blinded grading of steatohepatitis according to modified CRN criteria (Supplementary Table 3).[7] Liver fibrosis was measured by histologic grading of Masson's Trichrome staining or quantification of Picosirius Red positivity using ImageJ.

*Cell sorting*

Freshly perfused liver tissue was diced and digested with 850 mg/mL collagenase I, 700 mg/mL collagenase D, 1 mg/mL Dispase II and 100 ng/mL DNase I for 40 minutes in an orbital shaker at 37 °C and filtered through a 70 micron mesh filter. After red cell lysis, cells were resuspended in PBS supplemented with 2% FBS for flow cytometric analysis using a Sony MA900 cell sorter. The following antibodies (all from Biolegend) were used for multicolor flow cytometry: rat anti-mouse F4/80 FITC (BM8), rat anti-mouse Ly6C APC (HK1.4), rat anti-mouse CD3 PerCP-Cy5.5 (17A2), rat anti-human/mouse CD45R/B220 PerCP-Cy5.5 (RA3-6B2), mouse anti-mouse NK1.1 PerCP-Cy5.5 (PK136), rat anti-human/mouse CD11b APC-Cy7 (M1/70), rat anti-mouse Ly6G PE-Cy7 (1A8), mouse anti-mouse CD45.2 Pacific Blue (104), mouse anti-mouse CD45.1 BV510 (A20).

*RNA analysis*

RNA from freshly perfused mouse liver or sorted liver macrophages were extracted using Qiagen RNeasy Plus kit. Fragmentation, reverse transcription and cDNA library preparation with random hexamer primers were performed using standard Illumina protocols. Pooled libraries were sequenced using Illumina NovaSeq 6000. Sequenced reads were filtered to remove reads containing adapters, greater than 10% undetermined bases, or greater than 50% of bases with Q score less than or equal to 5. Transcript abundance estimates were generated using Salmon v1.2.1 and differentially expressed genes ($log_2FC > 0.58$, padj < 0.05) identified using R package *DESeq2*. Gene set enrichment analysis was carried out using GSEA v4.1.0 (Broad Institute). For sorted liver macrophages, gene sets in MSigDB C7 (filtered for macrophage-related signatures) and H (Hallmark) collections were analyzed. For unsorted liver transcripts, gene sets in MSigDB C2 (filtered for liver-related signatures) and H collections were analyzed, in addition to liver disease-specific gene sets extracted from published data.[8–10] P value < 0.05 and FDR < 0.15 were taken to be significant.

*Statistical analysis*

We tested for the association of CHIP status with prevalent chronic liver disease using logistic regression, with adjustment for age, sex, type 2 diabetes and smoking. We conducted an additional analysis with adjustment for alcohol consumption and body mass index. For incident chronic liver disease, we tested the association of carrying a CHIP variant with incident disease using Cox proportional hazards regression, with adjustment for age, sex, type 2 diabetes and smoking. Further adjustment for alcohol consumption and body mass index was also performed as a sensitivity analysis. To pool the association of CHIP with liver disease across cohorts, inverse variance weighted fixed effects meta-analysis was performed. Fixed effects meta-analysis was used given the lack of heterogeneity between estimates from different cohorts. To examine whether clone size may modify the association of CHIP status with liver disease, as has been reported for hematologic malignancy and cardiovascular disease,[5,11,12] we stratified patients by variant allele fraction ≥ 0.1 and < 0.1 and mutated gene. We tested for the association of CHIP status with liver fat (proton density fat fraction) and liver inflammation and fibrosis (proton cT1), we used logistic regression with adjustment for age and sex. Fatty liver was defined as fat fraction ≥ 5%. Fibrotic liver was defined as proton cT1 ≥ 795 ms.[3]

To assess the causality of the association of CHIP status with chronic liver disease, we performed a Mendelian randomization analysis. We examined the association of genetic predisposition to CHIP with cirrhosis risk. To increase statistical power to detect an effect, we used variants associated with CHIP status at a p-value of less than 0.0001. Pruning was performed to identify independent genetic variants using $R^2 < 0.01$ prior to analysis. The GWAS for the exposure (CHIP) consisted of a genome-wide association study of CHIP status in 52 studies in the Trans-omics for Precision Medicine (TOPMED). Among 97,691 individuals, 4,229 cases and 93,462 controls were analyzed. Single variant association for each variant with MAF > 0.1% and MAC > 20 was performed with SAIGE. Models were adjusted for age, sex and ten principal components of ancestry. The genetic variants identified exhibited strong association with CHIP (F-statistic = 156).

We tested the association of these variants with cirrhosis risk using summary statistics from a genome wide association study of 5770 cirrhosis cases and 487780 controls.[1] This genome wide association study analyzed all-cause cirrhosis - defined as hospitalization or death due to ICD10 codes K70.2, K70.3, K70.4, K74.0, K74.1, K74.2, K74.6, K76.6, or I85. Logistic regression as implemented in PLINK was used to test the association of genetic variants with all-cause cirrhosis in seven cohorts. Inverse variance weighted meta-analysis was used to pool estimates across all seven cohorts. All analyses were adjusted for age, sex and five principlan components of ancestry. Cohorts were of European ancestry. We used the Mendelian randomization with robust adjusted profile score method, which provides for control of type 1 error rate when using sub-genome wide significant genetic variants.[13] In sensitivity analyses, we also performed MR analysis using MR-PRESSO[14], MR-EGGER[15] and multivariate MR adjusted for smoking, body-mass index and type 2 diabetes[16–18]. No outliers were detected in the MR-PRESSO analysis. Full summary statistics for the cirrhosis GWAS are available for download at https://cvd.hugeamp.org/downloads.html. All statistical analyses were conducted using R version 3.5.

For mouse studies, pairwise comparisons were made using Student's t-test for parametric data and Mann Whitney U test for non-parametric data. Statistical analyses were performed in R version 3.5 or Graphpad Prism 8.3.1.
